# Climate factors and child health in Sweden: a nationwide whole-population study of 4.6 million school-aged children

**DOI:** 10.64898/2026.10.05.26364616

**Authors:** Shivang Pandey, Leonardo Olivetti, Isabel Johanna Walter, Max Wybrant, Luigi Filippo Brizzi, Francesca Cotta Ramusino, Pia Hardelid, Manuel Martellini O Nocentini, Stefan Döring, Carlo Torti, Maurizio Mazzoleni, Cecilia Magnusson, Gabriele Messori, Elena Raffetti

## Abstract

**Background:** Short-term climatic exposure conditions can trigger paediatric hospital admissions, but evidence remains limited on the health relevance of intraday temperature variability, humidity, and precipitation, and generally on which subgroups are most susceptible.

**Methods:** We conducted a nationwide case time-series study of Swedish children and adolescents aged 6–18 years, between 1987–2018 using linked register data. Associations between daily mean temperature, intraday temperature variability, relative humidity, and precipitation and inpatient hospitalisations were estimated with Poisson regression (individual fixed effects) combined with distributed lag non-linear models over lags 0–21 days. Outcomes included all-cause hospitalisation and cause-specific admissions. Effect modification was examined by sex, age group, socioeconomic indicators, region, and calendar periods.

**Findings:** Among 4,638,247 children and adolescents in the cohort, 1,070,702 (23·1%) experienced at least one hospitalisation, contributing 1,736,169 hospitalisation events. All-cause hospitalisation increased with higher mean temperature, with risk highest at hot extremes. Intraday temperature variability showed a U-shaped association, with higher risks at both lowest and highest exposure levels. Relative humidity showed no association with hospitalisation risk. Higher precipitation was associated with increased hospitalisation risk, but estimates at high rainfall levels were imprecise. Stratified analyses indicated modest differences by sex and socioeconomic indicators, but clearer heterogeneity by age and region, with the highest risks among younger children and in East Sweden; associations varied across calendar periods and were imprecise for most cause-specific outcomes. Sensitivity analyses mostly showed that the associations were robust.

**Interpretation:** Acute exposure to hot temperatures, both low and high intraday temperature variability, and higher levels of precipitation were associated with an increased risk of all-cause hospitalisation among children in Sweden, with clearer variation by age group and region than by sex or socioeconomic position. These findings support targeting paediatric climate-health preparedness and early-warning systems toward younger children and higher-risk regions such as eastern Sweden and extending such systems beyond mean temperature to include intraday temperature variability, a distinct, currently overlooked driver of risk that heat-focused systems are not designed to capture.

**Funding:** This research was funded by Formas the Swedish Research Council for Sustainable Development (grant no. 2023-01774 and 2022-01845) and the Swedish Research Council (2025-02804 and 2022-06599).

**Research in context:** *Evidence before this study:* We identified epidemiological studies examining climatic exposures and hospitalisation among children, published in any language from February 1, 2025, the final search date of the most recent systematic review, to June 26, 2026, in MEDLINE, Embase and Web of Science. Search terms combined paediatric populations and hospital admissions with climate exposures and distributed lag non-linear models (DLNM) (eg, (child OR paediatric*) AND (hospital* OR admission*) AND (temperature OR heat OR cold OR “diurnal temperature range” OR “temperature variability” OR intraday AND humidity OR precipitation OR rainfall) AND (DLNM OR “distributed lag” OR case-time series* OR case-crossover*). Previous studies have reported associations between ambient temperature and increased risk of paediatric morbidity, particularly for respiratory and infectious diseases. However, evidence is often limited to small study population, single healthcare settings or geographic areas, cause specific outcomes, aggregated data, and incomplete assessment of lagged effects and subgroup vulnerability. Analyses of different climatic exposures, such as temperature, humidity, and precipitation, remain rare.

*Added value of this study:* Our study used linked whole-population register data from more than 4·6 million school-aged children and adolescents aged 6–18 years in Sweden. This is one of the largest whole-population cohort studies of individual-level climate–health associations in children internationally, and the first such study in the Nordic countries to examine the short-term associations between climatic exposures and hospitalisation risk for children. We found that daily mean temperature and intraday temperature variability were each associated with all-cause hospitalisation, with higher risks at hot temperature extremes and at both low and high levels of within-day temperature variability. Relative humidity showed no association, whereas higher precipitation was associated with increased hospitalisation risk, although estimates became statistically weaker and more imprecise at the highest rainfall levels. The study has several important strengths: it examines different climatic exposures within a single analytical framework; it assesses heterogeneity by age, sex, socioeconomic indicators (parental education and household income), region, calendar period, and parental medical history, showing that risk was highest among younger children and in East Sweden; it evaluates lag-specific and cause-specific associations, showing that heat-related risk was concentrated within the first week of exposure while cause-specific estimates were less precise; and it uses individual-level adjustment for time-invariant characteristics through a within-person case time-series design, controlling for time-invariant individual characteristics and therefore strengthening causal interpretation.

*Implications of all the available evidence:* Paediatric hospitalisation risk rises in the days following temperature extremes and periods of high intraday temperature variability, with the largest effects among younger school-age children (6–13 years) and in East Sweden. This supports practical measures such as ensuring reliable access to hydration and shifting the timing of outdoor activity to cooler parts of the day, and, at the healthcare-system level, regionally targeted paediatric surge planning; for example, aligning staffing and emergency capacity with forecasted periods of extreme or highly variable temperature. The chief implication for scientific understanding is that within-day temperature variability represents a distinct dimension of climate-related risk not captured by mean or maximum temperature alone, suggesting heat-health frameworks and early-warning systems built on mean temperature capture only part of the risk. The U-shaped association, with increased risk under both very stable and highly variable conditions, indicates that temperature variability carries information beyond mean thermal load and warrants further investigation. Future research should test whether incorporating variability–metrics improves prediction of paediatric admissions and whether these patterns generalise beyond high-latitude settings.

## Introduction

Climate change is raising global temperatures and leading to more frequent weather extremes, posing major public health risks for current and future generations. The mean global temperature in 2023-2025 exceeded 1·5°C above the pre-industrial baseline, and Europe is currently warming at more than twice the global average.^1^ The health impacts are already evident, including 181,000 heat related deaths across Europe in the summers of 2022, 2023, and 2024.^2^ Cold spells continue to account for substantial morbidity and mortality and are projected to persist as a recurrent hazard in northern regions for the coming decades.^3^ Climate change also modifies other climatic exposures, including patterns of short-term temperature variability, humidity during warm periods, rainfall and floods, with potential impacts on population health.^4–6^

School-aged children are more vulnerable than working-age adults to non-optimal (moderate departures from the climatic variable value at which risk is lowest) and extreme weather conditions owing to their developing physiological systems and longer time spent outdoors.^7,8^ These climatic factors exert both direct and indirect impacts on child health.^9^ For example, extreme heat impairs thermoregulation and increases respiratory and gastrointestinal morbidity, while cold can trigger airway inflammation.^10,11^ Indirectly, extreme precipitation may limit access to care.^12^ Children in lower-income households are likely to bear a disproportionate burden through greater exposure, via housing and neighbourhood conditions such as limited access to thermally regulated indoor spaces, and greater vulnerability at comparable levels of exposure.^13^ This evidence, however, comes largely from single-site or single-city observational studies and selected populations, rather than whole-population data.

Climate-health associations remain limited for school-aged children, particularly in Sweden.^14^ Most studies have focused on older adults and single climatic exposures and relied on aggregated outcomes. They rarely examine how vulnerability is shaped by developmental stages, school year, geographic location, and socioeconomic factors.^13,15^ As a result, public-health agencies, schools, and health services lack the evidence needed to decide which climatic conditions warrant preparedness for children, beyond heat alone, and which groups of children to prioritise. To overcome these limitations, we linked whole-population Swedish health and administrative registers for children aged 6–18 years to high-resolution climate data to quantify associations of non-optimal and extreme temperature (mean and intraday variability), relative humidity, and precipitation with risk of hospitalisation. We modelled non-linear, lagged effects using a within-person design and assessed effect modification by age, sex, socioeconomic indicators, region, and calendar period.

## Methods

### Study Design and Population

We conducted a whole-population case time-series study using linked data from six Swedish national registers: the Total Population Register (RTB), the Longitudinal Integrated Database for Health Insurance and Labour Market Studies (LISA), and the National Education Register (UREG) from Statistics Sweden (SCB); and the Medical Birth Register (MFR), National Patient Register (NPR), and Cause of Death Register from the Swedish National Board of Health and Welfare (Socialstyrelsen). Children were linked to their parents using parental identifiers recorded in the Medical Birth Register; parental education, income, and medical history were derived from LISA, RTB, and the National Patient Register, respectively.

We defined a whole-population cohort of children and adolescents (hereafter, for brevity, “children”) aged 6–18 years in Sweden between 1 January 1987 and 31 December 2018 using the RTB (see Supplementary Methods for the rationale for this age range). Children entered the cohort from the start of the calendar year in which they turned six years, or from first registration in Sweden if later, and were followed until the end of the calendar year in which they turned 18 years, death, emigration/deregistration, or 31 December 2018, whichever occurred first. Because both cohort entry and exit were defined by calendar year rather than exact birthdate, the age distribution of the cohort was broadly consistent across the study period rather than skewed toward younger ages in earlier calendar years.

We applied a case time-series design.^16^ Analyses were restricted to individuals who experienced the outcome, with each individual serving as their own control on non-event days within the same calendar month. Children could be admitted to hospital more than once during follow-up. Because admissions occurring close together often represent readmission or continuation of the same illness episode rather than a new, independent event, we applied a 28-day ‘washout’ period: any hospitalisation occurring within 28 days of a previous one was excluded, and only admissions separated by at least 28 days were retained as distinct events. Each retained event was then analysed within its own person-month-year stratum, so that comparisons were made only between days within the same individual and calendar month, which accounts for within-person clustering by design (see Supplementary Methods for implementation details). Climate exposure on the event day was compared with exposure on selected non-event days for the same individual, enabling estimation of short-term effects and yielding a within-person design that controls for all time-invariant confounders. The case time-series analysis rests on the assumptions that: i) time-varying confounding within each person-month-year comparison window is adequately controlled (via day-of-week and week-of-year); ii) the occurrence of an event does not alter the probability of a subsequent event; iii) exposure measurement error is non-differential with respect to event timing and iv) each child’s registered parish or district approximates where they were actually located on the days of exposure, so that climate values assigned from that location approximate their true exposure.

Data were accessed in the National Academic Infrastructure for Supercomputing in Sweden. The analysis was performed according to a prespecified protocol. The phenotypes and associated codes are available at [Supplementary Methods and *github.com/shivangpandey31/climate-school-children-health-sweden*]. The study is reported in agreement with the RECORD and STROBE statements for observational studies using routinely collected health data.

### Variables of the study

#### Outcome Variable

The primary outcome was all-cause hospitalisation. As secondary outcomes, we included cause-specific hospitalisation for injury and poisoning, respiratory diseases, digestive diseases, and mental disorders, selected as the most frequent ICD-chapter groupings among hospitalisation events within the age range. Cause-specific outcomes were defined using the primary diagnosis code. Full details of the code list applied, and information on admission type, are provided in Supplementary Methods.

#### Exposure

Daily environmental exposures were derived by linking individuals’ annual parish/district of residence (from SCB registers; see Supplementary Methods for parish size and population characteristics) to harmonised administrative boundaries, with all historical units reconciled to a consistent 2015 parish geography using shapefiles from the Swedish national mapping, cadastral and land registration authority (Lantmäteriet). Gridded climate data were obtained from the Swedish Meteorological and Hydrological Institute (SMHI) GridClim^17^ (2·5×2·5 km; 1961–2018) and recast to parish-level, population-weighted daily means, which were then assigned to individuals. We examined four exposures: daily mean ambient temperature (DAT), daily mean relative humidity (RH), daily total precipitation (PR, converted to mm/day), and intraday temperature variability (ITV), defined from 6-hourly temperatures (00:00, 06:00, 12:00, 18:00) as the sum of absolute successive within-day changes (°C/day). See [Supplementary Methods] for more information on the climate data.

#### Effect Modifiers

We examined effect modification by sex (Female/Male) as recorded in the Total Population Register (legal sex); age group (6–9, 10–13, 14–18 years; aligned with the Swedish education system^18^), parental education (highest completed level between both parents, measured one year before the child’s birth: primary school, upper secondary, university); household disposable income (family disposable income, measured one year before the child’s birth and divided into population quintiles); calendar period based on change in education policy and economic context over time (1987–1993, 1994–2005, 2006–2018);^19^ geographic region (based on residence; NUTS1 regions: East Sweden , South Sweden, North Sweden);^20^ and parental medical history defined as any parental diagnosis, from a pre-specified list of ICD codes recorded in the National Patient Register before the child’s birth (full code list in Supplementary Methods). In addition to NUTS1 region, we examined spatial variation at the county level (län) which generally Swedish health care regions, also derived from each individual’s registered residence on the population register. County-specific IRRs were estimated at selected values spanning the range of each exposure and are presented as maps (Figure 4; relative humidity in Supplementary Figure S14). See the Supplementary Methods for more information on the description of cohort.

### Statistical Analysis

#### Statistical Model

We applied a case time-series analytical framework employing distributed lag non-linear models (DLNM) to estimate associations between climate exposures and hospitalisation outcomes. ^21,22^ Incidence rate ratios (IRRs) were estimated across the full observed exposure distributions of temperature, intraday temperature variability, relative humidity, and precipitation, centred at the median of each variable.

For each exposure, we constructed a cross-basis using natural cubic spline functions for the exposure-response dimension. Internal knots were placed at the 10^th^, 75^th^ and 90^th^ percentiles of the exposure distribution except for precipitation. There, boundary knots were set at the 37^th^ (around 0·2 mm/day) and 99^th^ (around 21·8 mm/day) percentiles with internal knots at 50^th^, 75^th^ and 90^th^ percentiles to stabilize estimates in the tails of the distribution given the high frequency of zero-rain days and the long right tail of extreme precipitation values. The lag-response dimension was modelled within the same DLNM *crossbasis* over 0–21 days. Cumulative exposure-response associations were obtained using *crosspred*. In practical terms, an IRR at a given exposure level reflects the cumulative change in hospitalisation risk associated with a single day at that exposure, relative to a single day at the median exposure level, summed over that day and the following 21 days of the lag window (i.e., combining the immediate effect with delayed contributions to risk).

We estimated the short-term effect of climatic exposures on daily hospitalisations using a fixed-effect Poisson regression with each stratum represented by individuals as person-month events. Let, *Y*_*i,t*_, denote the outcome indicator for each of the *i* = 1, *…* , *n* individuals on day *t*. The model is specified as

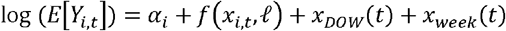

where *α*_*i*_ are individual-specific intercepts and *f*(·) is the DLNM *crossbasis* capturing non-linear and lagged temperature responses over the lag period *L =* 0, … , 21. To control for time-varying confounding, we included time-varying control terms *x*_*a*_ (*t*) for day of week and week of year, modelled as categorical indicators. Week of year, rather than fixed calendar date, was used for two reasons: school-holiday periods (eg, Midsommar and other school breaks) shift from year to year by International Organisation for Standardisation (ISO) week, and hospital staffing levels are also known to vary by week of year (eg, during common vacation periods), independently of climate exposure.

We additionally report the minimum morbidity value (MMV), the exposure level at which hospitalisation risk was lowest, sometimes termed the optimal value as a contrast point in stratified analyses; non-optimal exposure refers to any departure from this value, and extremes to values below the 1st or above the 99th percentile.

Effect modification was assessed formally by adding interaction terms between the cross-basis and each modifier to the main model and testing all interaction coefficients jointly with a Wald χ^2^ test.

We conducted six sensitivity analyses. First, we repeated the analysis using alternative lag structures of 3, 7, 14, 21, and 28 days, to evaluate how the choice of exposure window influenced the estimated climate– hospitalisation associations. Second, we compared DLNM specifications with and without boundary knots in the cross-basis, to evaluate sensitivity to tail modelling beyond extreme percentiles. Third, we excluded summer and Christmas breaks and, separately, stratified by school holiday status (see Supplementary), to assess potential bias from holiday-related changes in hospitalisation patterns. Fourth, we replicated the analysis using a time-stratified case-crossover design, selecting control days falling on the same day of the week within the same calendar month as the event day, to assess consistency with the case time-series result.^23,24^ Fifth, we restricted the analysis to each child’s first recorded hospitalisation during follow-up, to assess whether repeated admissions of the same child, which may not be independent of one another, influenced the results. Sixth, we compared hot (April–September) and cold (October–March) seasons, with spline knots recomputed separately within each season using the same percentile-based approach described above.

Python 3·11 was used to construct the data processing pipeline and creating figures using *pandas* (2·3·2) and *matplotlib* (3·9·2), while statistical analyses were carried out in R 4·3·1 using the *dlnm* (2·4·7), and *gnm* (1.1·5) packages.

## Results

### Descriptive statistics

Of the 4,638,247 children in the cohort, 1,070,702 (23·1%) experienced at least one inpatient hospitalisation during the study period (Table 1). In total, 2,029,276 hospitalisation events were identified, of which 1,736,169 remained after applying the 28-day washout. Among these retained events, 411,024 (23·7%) were for injury and poisoning, 191,997 (11·1%) for respiratory diseases, 149,150 (8·6%) for digestive diseases, and 107,890 (6·2%) for mental disorders. The remaining 876,108 events (50·5%) fell outside these cause-specific groupings; the largest share of these (233,304 events, 13·4%) were classified as symptoms and ill-defined conditions (ICD-9 780–799; ICD-10 R00–R99).

**Table 1.** Baseline characteristics of the cohort, by hospitalisation status. Values are numbers of children (%)^1^.

| Characteristics | Total (n=4,638,247) N (%) | Hospitalised<br>(n=1,070,702) N (%) | Non-hospitalised<br>(n=3,567,545) N (%) |
| --- | --- | --- | --- |
| **Sex** |  |  |  |
| Male | 2,390,845 (51.55%) | 571,737 (53.40%) | 1,819,108 (50.99%) |
| Female | 2,247,402 (48.45%) | 498,965 (46.60%) | 1,748,437 (49.01%) |
| **Parental education** |  |  |  |
| Primary School | 491,795 (10.60%) | 121,991 (11.39%) | 369,804 (10.37%) |
| Upper secondary | 2,059,955 (44.41%) | 536,059 (50.07%) | 1,523,896 (42.72%) |
| University | 1,958,203 (42.22%) | 390,176 (36.44%) | 1,568,027 (43.95%) |
| Missing | 128,294 (2.77%) | 22,476 (2.10%) | 105,818 (2.97%) |
| **Income quintile** |  |  |  |
| 1 (Lowest) | 879,199 (18.96%) | 186,069 (17.38%) | 693,130 (19.43%) |
| 2 | 864,157 (18.63%) | 206,167 (19.26%) | 657,990 (18.44%) |
| 3 | 882,502 (19.03%) | 214,825 (20.06%) | 667,677 (18.72%) |
| 4 | 919,755 (19.83%) | 219,413 (20.49%) | 700,342 (19.63%) |
| 5 (Highest) | 991,101 (21.37%) | 223,903 (20.91%) | 767,198 (21.50%) |
| Missing | 101,533 (2.19%) | 20,325 (1.90%) | 81,208 (2.28%) |
| **Parental medical history** |  |  |  |
| With | 1,754,527 (37.83%) | 440,258 (41.12%) | 1,314,269 (36.84%) |
| Without | 2,791,953 (60.19%) | 611,923 (57.15%) | 2,180,030 (61.11%) |
| Missing | 91,767 (1.98%) | 18,521 (1.73%) | 73,246 (2.05%) |

Figure 1 shows the cumulative exposure-response associations with all-cause hospitalisation over 0–21 lag days. Risk increased with higher daily mean temperature: cumulated over the day of exposure and the following 21 days, the rate of hospitalisation associated with a day at 24·4 °C (99th percentile) was 22% higher than for a day at the median temperature (6·4 °C). By contrast, risk was reduced at low temperatures, with an IRR of 0·91 (0·88−0·95) at -14·7°C (1st percentile) relative to the median. Intraday temperature variability showed a U-shaped association, with higher risks at both low (IRR = 1·06 (1·02–1·11) at 1·3 °C/day – 1^st^ percentile) and high variability (IRR = 1·08 (1·03–1·13) at 28·6 °C/day – 99^th^ percentile). Precipitation showed a non-linear association, with risk increasing above approximately 20 mm/day. Relative humidity was not associated with hospitalisation, with estimates remaining close to the null across the exposure range. Seasonal stratification suggested stronger associations for intraday temperature variability during October-March (Supplementary Figure S7). Unless otherwise stated, all incidence rate ratios reported below represent cumulative risk over the full 0–21 day lag window.

**Figure 1:**
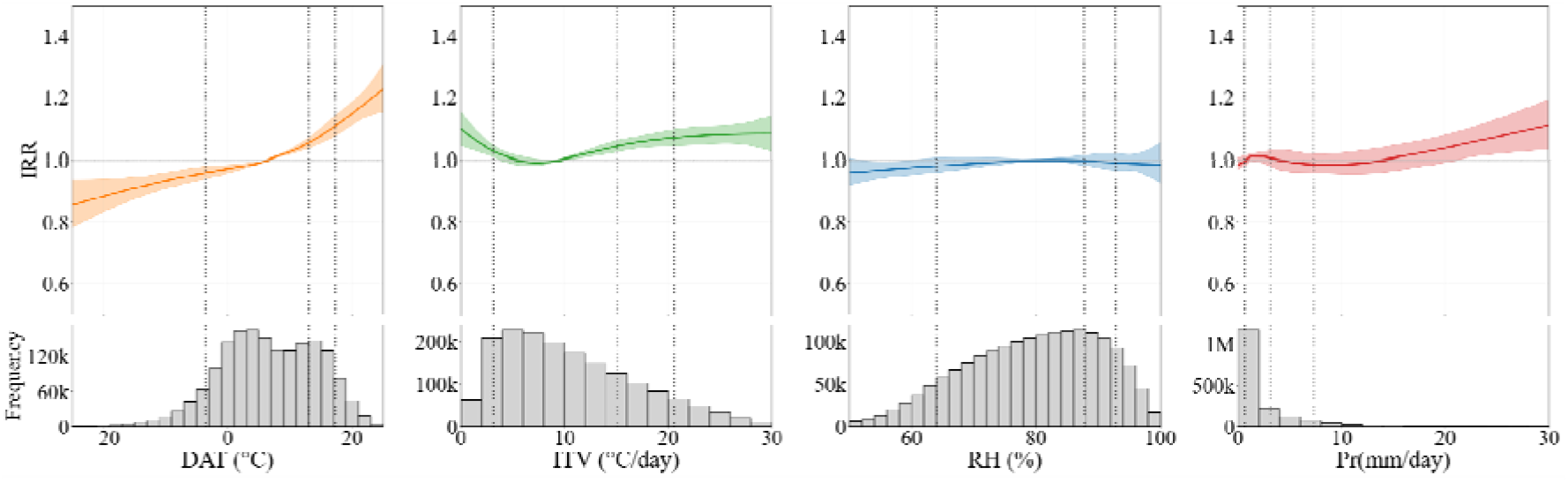
Overall cumulative exposure-response associations between climate exposures and all-cause hospitalisations over 21 lag-days for Daily Average Temperature (°C), Intraday Temperature Variability (°C/day), Relative Humidity (%) and Daily Precipitation (mm/day) (from left to right). Curves show incidence rate ratios or IRRs (solid lines) and 95% confidence intervals (shaded areas); estimates are centred at the median of each exposure. We have histogram and vertical dotted line shows where we have put not for each exposure

Figure 2 shows a contour plot of exposure by lag day, with IRR on the z-axis. Elevated risk at high daily mean temperature was concentrated at short-to-mid lags (∼ days 1–7), while reduced risk at low temperature was concentrated at short lags only (∼ days 1–5) and did not persist to longer lags. High intraday temperature variability showed a similar pattern of elevated risk concentrated at short-to-mid lags; the association at low variability was weaker in magnitude and less consistent across the lag period. Precipitation showed a weak, positive association that varied little across the lag period. Further analyses for daily mean temperature, intraday temperature variability, and precipitation are presented in the main text below. As relative humidity showed no clear association, further results for this exposure are presented in Supplementary Figure S15 and S16.

**Figure 2:**
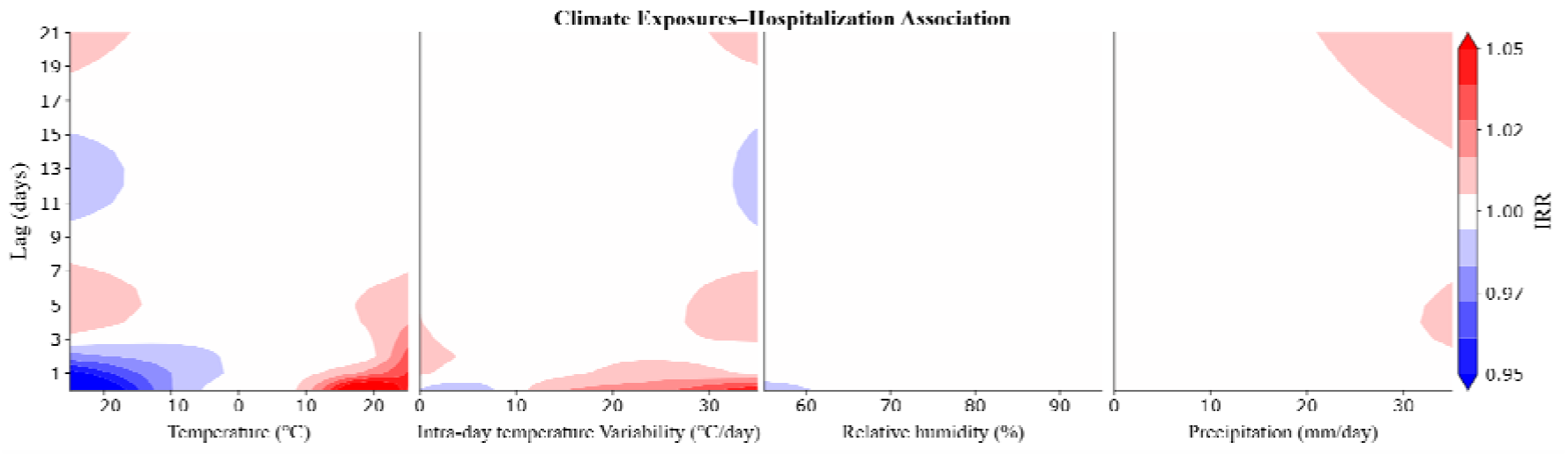
Contour plots for Daily Average Temperature (°C), Intraday Temperature Variability (°C/day), Relative Humidity (%) and Daily Precipitation (mm/day) (from left to right)

For cause-specific analysis, injury and poisoning shows increased risk with higher temperature and temperature variability but other estimates were generally close to the null or imprecise with wide confidence intervals and no clear exposure–response patterns (Supplementary Figure S10).

Figure 3 shows the effect modification by those factors for which curves between strata were the most different (age, period, and region). Associations at high mean temperature and temperature variability were stronger in younger children (6–9 and 10–13 years); at lower temperatures, the pattern was similar with lowest risk associated with the oldest age group (14–18 years). Period differences were most evident for daily mean temperature, with a steeper rise at higher temperatures in the most recent period. For precipitation, higher rainfall showed stronger positive associations during 2006–2018, although estimates at the upper tail remained uncertain.

**Figure 3:**
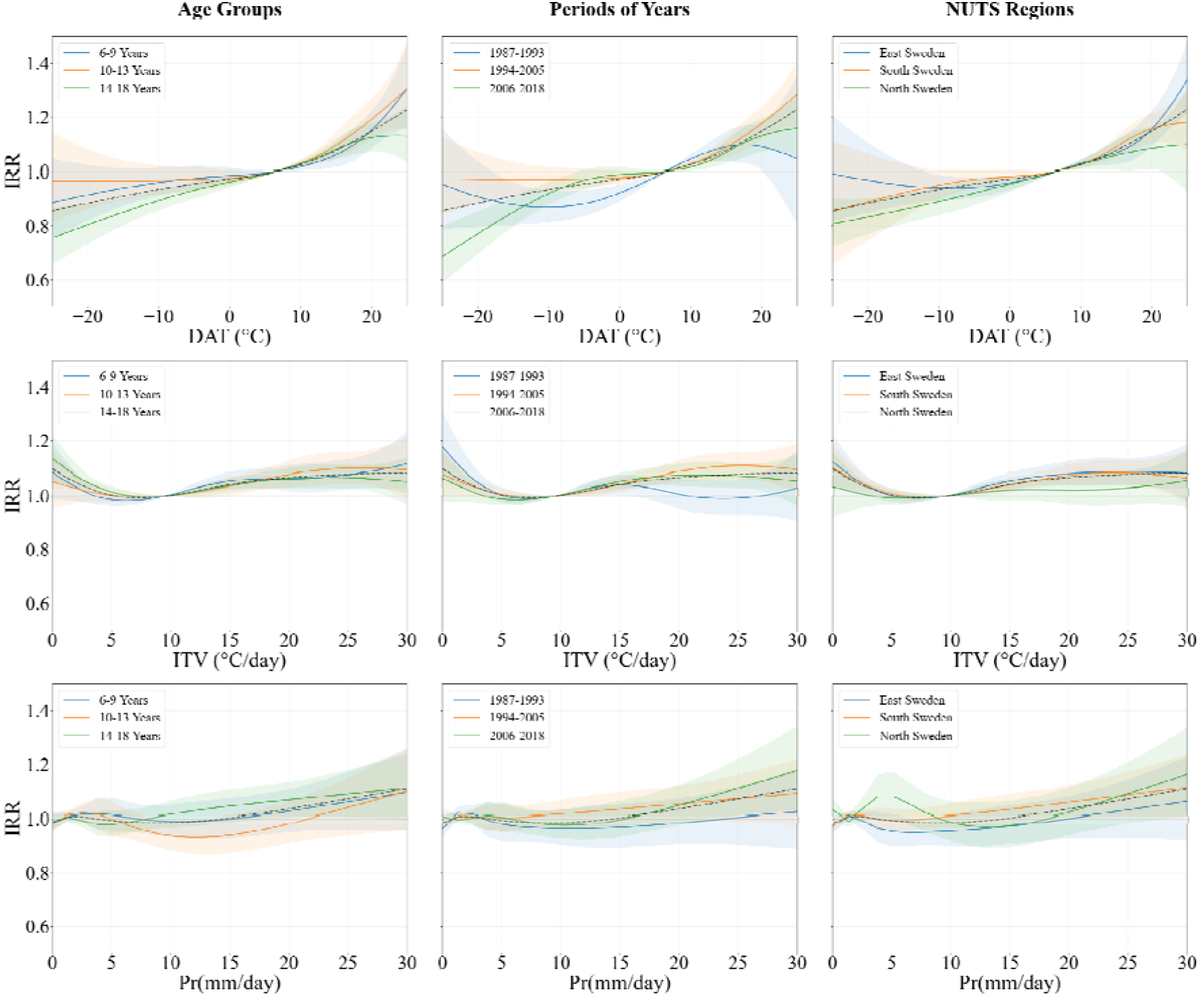
Stratified exposure-response associations for all-cause hospitalisations. IRRs are shown for Daily Average Temperature (°C), Intraday Temperature Variability (°C/day) and Precipitation (mm/day) (top to bottom) by age group, calendar period, and region; the dashed grey line shows the corresponding overall, unstratified association for comparison

Regional variation was concentrated at the extremes of the exposure distributions. At high daily mean temperatures, hospitalisation risk increased most steeply in East Sweden, followed by South Sweden, with the smallest increase in North Sweden. At low temperatures, IRRs were below the reference in all regions and lowest in North Sweden. Associations with intraday temperature variability were similar across regions at low-to-moderate values, whereas at high variability, risk increased more clearly in East and South Sweden than in North Sweden. For precipitation, the clearest regional signal was observed in North Sweden, where hospitalisation risk increased more strongly at higher rainfall levels than in East or South Sweden. We also examined risk stratified by individual counties, Figure 4, and observed similar spatial patterns: elevated risk at 25°C daily mean temperature in several central and eastern areas, at very low variability in a small number of southern counties, and at very high variability in central, eastern, and southern counties.

**Figure 4:**
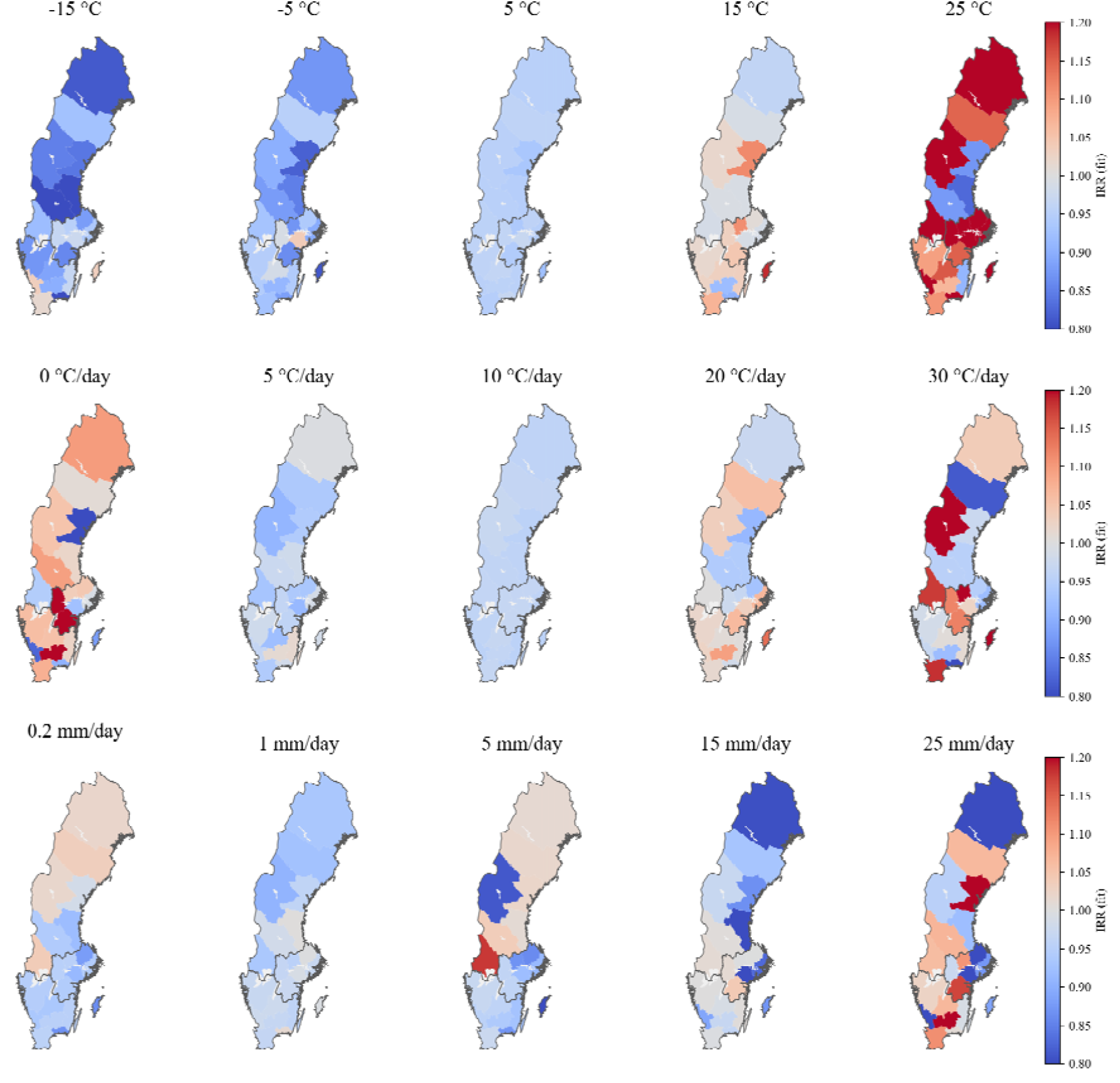
Incidence rate ratios (IRR) by Swedish county at selected exposure levels of daily average temperature (°C), intraday temperature variability (°C/day), and precipitation (mm/day) (top to bottom), relative to the median of each exposure (6·37°C, 9·8°C/day, and 0·65 mm/day, respectively).

For all other effect modifiers (sex, and parental income quintile, education and medical history), stratified curves were largely superimposable (Supplementary Figure S12). To examine whether risk between these strata nevertheless varied at climate extremes, we visualised contrasts at the 1st (37th for precipitation, 0·2 mm/day) and 99th percentiles of climate variables compared to the median (Figure 5, with likewise results for region, period and age in Supplementary Figure S13). At low temperatures, risk was slightly higher in males than females. Risk at high temperatures did not differ consistently by parental income (interaction p = 0·34). At low temperatures, IRRs followed a gradient from the lowest-income group (0·82, 95% CI 0·75–0·90) to the highest (0·98, 0·90–1·07), though this was not statistically significant. Children of parents with only primary education had higher IRRs for both mean temperature and intraday variability (Wald test for interaction, p = 0·053 and 0·044). Precipitation-related risks varied by income without a consistent gradient. Estimates were similar for children with and without a parental medical history.

**Figure 5:**
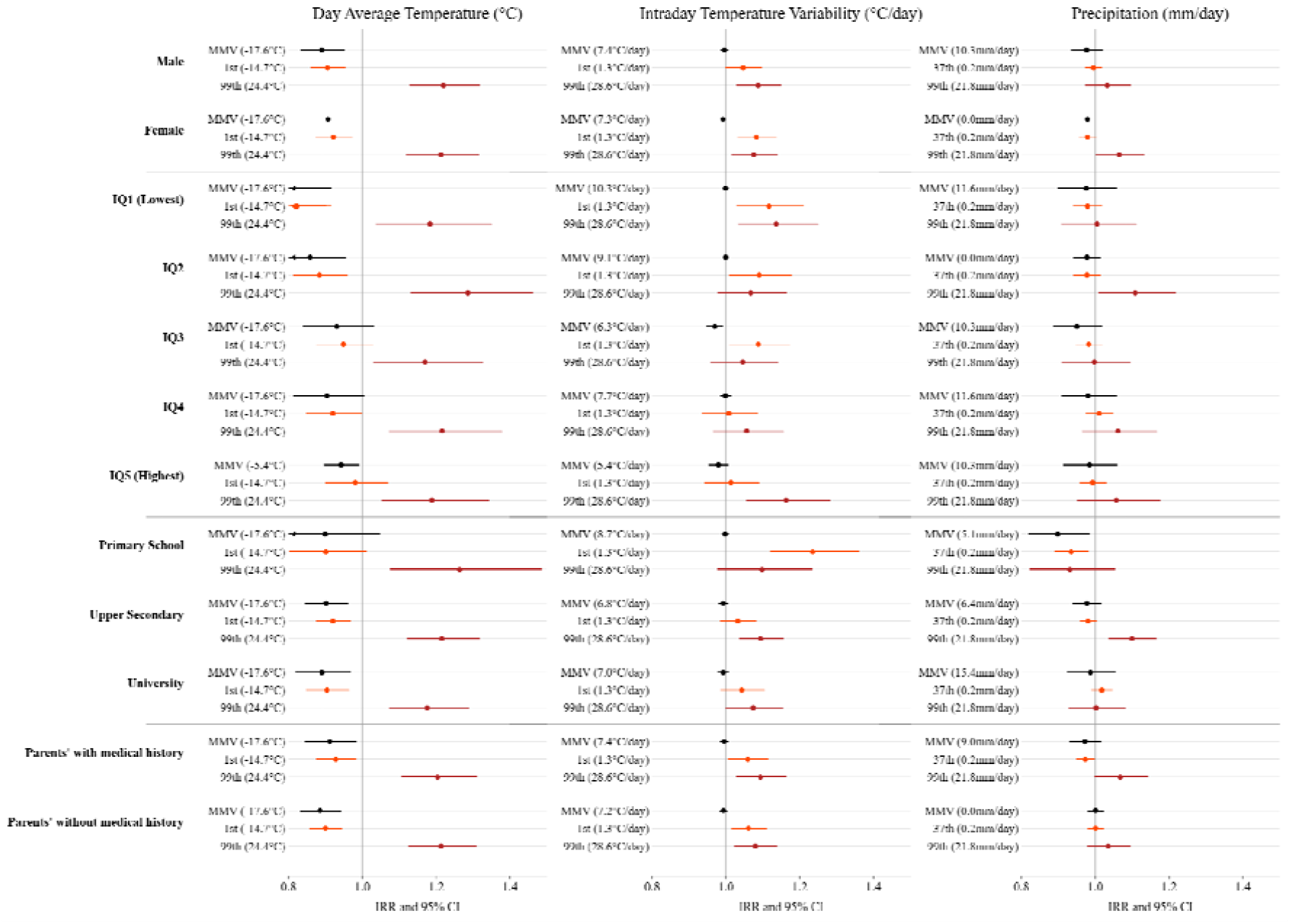
Stratified exposure-response associations for all-cause hospitalisations. IRRs are shown by sex, age group, parental education and individuals with parental medical history for 1^st^ (37^th^ for precipitation), 99^th^ percentile and minimum morbidity value (MMV), of Daily Average Temperature (°C), Intraday Temperature Variability (°C/day) and Precipitation (mm/day).

Sensitivity analyses showed similar patterns throughout: the shape and direction of the exposure–response curves were preserved, with differences confined to the magnitude of estimates and to the tails of the exposure distributions. First, shorter lag windows attenuated cumulative associations. Second, boundary knots added little information because tail observations were sparse; this was most relevant for precipitation, where many zero-rain days and few extreme rainfall events made tail estimates unstable. Third, results were similar when school holidays were excluded or stratified by, except for daily mean temperature. During school terms, extreme cold was associated with higher risk. This suggests that the lower risk on cold days in the main analysis largely reflects fewer admissions during the Christmas break, which coincides with some of the coldest weeks of the year. Heat estimates outside holidays were imprecise. Fourth, the time-stratified case-crossover design produced results similar to the main case time-series analysis. Fifth, restricting to each child’s first hospitalisation gave similar results, with a slightly stronger association at high daily mean temperature (see Supplementary Figure S11 for sensitivity analysis plots). Sixth, seasonal stratification suggested stronger associations for intraday temperature variability during October–March than April–September (Supplementary Figure S7).

## Discussion

Our study has shown that high daily mean temperature, both low and high intraday temperature variability, and higher precipitation are associated with an increased risk of hospitalisation among 1·07 million Swedish children aged 6–18 years; while no association is observed for relative humidity.

Several mechanisms could plausibly link these exposures to paediatric hospitalisation, and the relevant pathway likely depends on the reason for admission. Injuries, about a quarter of admissions, showed the clearest association with heat and high intraday temperature variability, which is more plausibly behavioural than physiological: warm weather brings more outdoor activity, swimming, travel, and holiday activities, and with them more opportunity for injury. For medical conditions, high temperatures may act through impaired thermoregulation, dehydration, sleep disruption, and exacerbation of underlying conditions.^9^ Injuries, however, accounted for about a quarter of admissions and showed the strongest associations with both mean temperature and intraday variability. At high intraday temperature variability, rapid within-day changes may challenge thermoregulation and leave less time to adapt clothing, activity, hydration, and movement between indoor and outdoor environments. The association at very low variability is harder to explain. Days with very low variability occurred mainly in winter, so this association may reflect other conditions that accompany such days rather than variability itself. Very stable days are meteorologically distinctive. In Sweden, they occur during winter inversions and overcast conditions, but also during warm, cloudy periods in July. In winter, children spend more time indoors, favouring respiratory infections; in summer, they spend more time outdoors and travelling, which increases exposure to injuries. The low-variability association may therefore reflect seasonal conditions rather than stability itself. It was driven mainly by respiratory and digestive admissions, consistent with greater infection spread during cold, stable winter conditions, whereas the high-variability association was driven by injuries. Higher precipitation may increase risk through infection-related pathways in children.^12^ The timing of these associations pointed to short-term responses: risk at both high mean temperature (≥24°C, the 99th percentile) and high variability was concentrated within approximately the first week after exposure. Risk was instead lower on cold days, and this reduction occurred mainly in the first few days after exposure. deferred care-seeking during cold weather, when travel is difficult and non-urgent visits may be postponed, is a plausible contributor. This is consistent with the reduction being concentrated in symptoms and ill-defined conditions and in mental disorders rather than in injuries. These two groups may partly overlap, as admissions coded as symptoms in children are often related to mental health conditions. Because the reasons for hospital admission differ between children and adults, so may their associations with temperature. Findings from adult or all-age studies, which are dominated by adults, therefore cannot be generalised to children. Cause-specific patterns varied. Hospitalisation risk for injury and poisoning increased with both higher temperature and greater temperature variability, plausibly through increased outdoor activity, risk-taking behaviour, dehydration, fatigue, and reduced concentration during hot or rapidly changing weather. Associations for respiratory and digestive diseases were positive but less precise.

Associations differed more clearly by age and region than by sex or socioeconomic position. Younger children had higher risks at high mean temperature and high intraday temperature variability, possibly reflecting differences in thermoregulation, activity, and time spent outdoors. Heat-related risk was greatest in East Sweden and weakest in North Sweden, while high intraday temperature variability showed stronger associations in East and South Sweden. East Sweden includes Stockholm, with urban heat islands and a diverse population with varying healthcare-seeking patterns, and admission thresholds may differ between regions. These factors, together with differences in housing and local climate adaptation^7,14^, may contribute to the regional patterns. Direct comparison with previous studies is difficult, as paediatric evidence has largely focused on single outcomes or settings,^11,28–30^ and few studies have examined multiple dimensions of susceptibility within one population. Differences by sex were small and, while there were modest differences by socioeconomic status, and those by socioeconomic position modest, with no consistent income gradient in heat-related risk, in contrast to settings with more unequal access to cooling and care,^31,32^ which may reflect Sweden having less social inequality and universal healthcare access. Associations differed across calendar periods, with little evidence of heat-related risk in 1987–1993 and clearer associations thereafter, although estimates for the earliest period were imprecise. These periods span substantial changes in Swedish society, including the economic crisis of the early 1990s, reforms to the education system, and a marked rise in wealth inequality from the 1990s onwards. Our design cannot identify which of these, if any, underlies the change, and period differences may also reflect changes in hospital admission practices and diagnostic coding over time. Heat-related risk nevertheless remained evident in the most recent period (IRR 1·16, 95% CI 1·06–1·26), indicating that it has not diminished despite three decades of social and health-system change.

Our study has several key strengths. The within-person case time-series design with individual fixed effects controls by construction for all time-invariant characteristics, and we additionally adjusted for time-varying factors including day-of-week and seasonality. Distributed lag non-linear models allowed us to capture non-linear exposure–response relationships and effects distributed across a 21-day lag window. Exposure assignment was strengthened in two ways: historical administrative boundaries were harmonised to a consistent 2015 geography to improve comparability over time, and population-weighted parish-level exposures were used to reduce misclassification arising from spatial aggregation. Finally, the nationwide whole-population cohort minimised selection bias and loss to follow-up, and provided the statistical power needed for precise estimation and for stratified analyses across Sweden.

Our study also has potential limitations. First, exposure assignment used registered district/parish of residence assumes children lived where they were registered; any mismatch may introduce non-differential misclassification and bias estimates toward the null. Second, although we controlled for seasonality (week of year), residual time-varying confounders (eg, circulating pathogens, behavioural changes) cannot be excluded. Third, estimates at the extremes particularly for heavy precipitation were imprecise, because such events were rare. Fourth, assessing compound exposures (notably temperature–humidity interactions to better capture heat stress)^4^ would be valuable, but joint DLNM modelling is not currently computationally feasible at the whole population scale, even with high-performance computing resources. Scalable approaches that can model multi-exposure, non-linear and lagged relationships such as machine learning or deep learning frameworks may offer a practical alternative for future work. ^33,34^

Heat–health action plans in Sweden and elsewhere in Europe are designed mainly around adults, and particularly older adults. Sweden’s framework consists of SMHI issued heat warnings, and Public Health Agency of Sweden (*Folkhälsomyndigheten)* provided guidance for municipal and regional heat-wave action plans. While young children are recognised as a vulnerable group,^35^ our findings suggest two changes. Warnings could take account of within-day temperature variability as well as mean temperature, and thresholds and paediatric service capacity could be set by region and age group rather than applied uniformly across the country. Swedish preschools have been shown to reach high indoor as well as outdoor temperatures, with little capacity to manage heat, so advice that assumes indoor spaces are cooler may not hold; ensuring children have reliable access to water, and moving outdoor activity to cooler parts of the day, are safer measures.^7,14^

High daily mean temperature and both low and high intraday temperature variability were consistently associated with increased short-term hospitalisation risk among school-aged children in Sweden. Children’s risk profiles differ from those established in adults and show no sign of diminishing over time. Climate–health preparedness that treats children as a distinct group, and that looks beyond mean temperature alone, is thus needed.

### Data sharing

The individual-level register data cannot be shared by the authors under Swedish law (Public Access to Information and Secrecy Act) and the General Data Protection Regulation (GDPR). Researchers can apply for the same data from Statistics Sweden and the Swedish National Board of Health and Welfare, subject to ethical approval by the Swedish Ethical Review Authority and the agencies’ review. Climate data are publicly available from SMHI (GridClim). The analysis code, code lists, and a data dictionary are available with publication at *github*.*com/shivangpandey31/climate-school-children-health-sweden*.

## Data Availability

The data analyzed in this study are derived from Swedish national registers managed by the National Board of Health and Welfare (Socialstyrelsen) and Statistics Sweden (SCB). Due to strict legal restrictions under Swedish legislation and GDPR regarding personal privacy and confidentiality, the dataset cannot be made publicly available or shared directly by the authors. Researchers interested in accessing these data for replication purposes must apply for ethical approval from the Swedish Ethical Review Authority (Etikprõvningsmyndigheten) and submit a formal data request directly to the respective managing authorities. Data access requests and procedures can be initiated via Socialstyrelsen and Statistics Sweden.

## Acknowledgment

This research was funded by Formas the Swedish Research Council for Sustainable Development (grant no. 2023-01774 and 2022-01845) and the Swedish Research Council (2025-02804 and 2022-06599).

The computations and data handling were enabled by resources in project sens2025030 provided by the National Academic Infrastructure for Supercomputing in Sweden (NAISS) at UPPMAX, funded by the Swedish Research Council through grant agreement no. 2022-06725.

## Ethical Approval

The Swedish Ethical Review Authority approved studies (reference 2020-05516, 2022-05648-02 and 2022-02626-01) to access de-identified, whole-population registry data, collected without individual consent from routine healthcare records and administrative registers.

## Contributors

SP and ER conceived and designed the study. ER and CM obtained the data and ethical approvals. SP and LO did the data curation. SP, LO, and MMON did the statistical analysis, with input from ER and GM; IJW contributed to the analysis code. SP and MMON accessed and verified the underlying data. ER, MM, and GM contributed expertise on the climate exposure data and their linkage. LFB, FCR, IJW, CM, and ER contributed to the ICD code lists and the classification of hospitalisations. SP, LO, IJW, MMON, and ER contributed to the methodology and the interpretation of the results. MW, PH, SD, and CT contributed to the interpretation of the findings and critically revised the manuscript. SP wrote the first draft of the manuscript. ER, MM, CM, and GM supervised the study. All authors reviewed and edited the manuscript, approved the final version, and had final responsibility for the decision to submit for publication.

## Declaration of use of Generative AI

During the preparation of this work, the author(s) used for Claude language support, structural feedback, and refinement of selected sections of the text. It was also used to help clarify concepts and improve phrasing. GitHub Copilot and Claude Code were used for programming support during data analysis and code development. The author(s) reviewed and edited the output as needed and take full responsibility for the content of the published article.

## Footnotes

1 All characteristics were measured before the child’s birth and are fixed for each child, so each child is counted once. Characteristics of hospitalisation events (n = 1,736,169), including cause of admission, are shown in Supplementary Table S1; children may contribute more than one event.

## References

1 Copernicus Climate Change Service. Copernicus: 2025 was the third hottest year on record. 2026; published online Jan. https://climate.copernicus.eu/copernicus-2025-was-third-hottest-year-record.

2 Janoš T, Quijal-Zamorano M, Shartova N, et al. Heat-related mortality in Europe during 2024 and health emergency forecasting to reduce preventable deaths. Nature Medicine 2025 31:12 2025; 31: 4065–74.

3 Gao Y, Huang W, Zhao Q, et al. Global, regional, and national burden of mortality associated with cold spells during 2000–19: a three-stage modelling study. Lancet Planet Health 2024; 8: e108–16.

4 Baldwin JW, Benmarhnia T, Ebi KL, Jay O, Lutsko NJ, Vanos JK. Humidity’s Role in Heat-Related Health Outcomes: A Heated Debate. Environ Health Perspect 2023; 131. DOI:10.1289/EHP11807.

5 Xu Z, Huang F, Liu Q, Fu C. Global pattern of historical and future changes in rapid temperature variability. Environmental Research Letters 2020; 15: 124073.

6 Liu Q, Fu C, Xu Z, Ding A. Global warming intensifies extreme day-to-day temperature changes in mid–low latitudes. Nature Climate Change 2025 16:1 2025; 16: 69–76.

7 Wallenberg N, Rayner D, Lindberg F, Thorsson S. Present and future heat stress of preschoolers in five Swedish cities. Clim Risk Manag 2023; 40: 100508.

8 Helldén D, Andersson C, Nilsson M, Ebi KL, Friberg P, Alfvén T. Climate change and child health: a scoping review and an expanded conceptual framework. Lancet Planet Health 2021; 5: e164–75.

9 Cousins S. Extreme weather events and child health. Lancet Child Adolesc Health 2019; 3: 70–1.

10 Schapiro LH, McShane MA, Marwah HK, Callaghan ME, Neudecker ML. Impact of extreme heat and heatwaves on children’s health: A scoping review. The Journal of Climate Change and Health 2024; 19: 100335.

11 Niu Z, Fang X, Sun J, et al. Exposure to Anthropogenic Heat Was Associated with the Increased Risks of Respiratory Allergy Symptoms in Preschool Children: A New Insight into the Urban Heat Island Effect. Environ Sci Technol 2025; 59: 12060–73.

12 Lai H, Hales S, Woodward A, et al. Effects of heavy rainfall on waterborne disease hospitalizations among young children in wet and dry areas of New Zealand. Environ Int 2020; 145: 106136.

13 Stowell JD, Sun Y, Spangler KR, et al. Warm-season temperatures and emergency department visits among children with health insurance. Environmental Research, Health 2022; 1: 015002.

14 Wallenberg N, Lindberg F, Thorsson S, et al. The effects of warm weather on children’s outdoor heat stress and physical activity in a preschool yard in Gothenburg, Sweden. Int J Biometeorol 2023; 67: 1927.

15 Wennman H, Partonen T. Impacts of temperature and solar radiation changes in northern Europe on key population health behaviors: a scoping review of reviews. Scand J Public Health 2023; 53: 184.

16 Gasparrini A. The Case Time Series Design. Epidemiology 2021; 32: 829–37.

17 Andersson S, Bärring L, Landelius T, Samuelsson P, Schimanke S. SMHI Gridded Climatology. 2021.

18 Utbildningsdepartementet. Skollag. Sveriges Riksdag, 2010 https://www.riksdagen.se/sv/dokument-lagar/dokument/svensk-forfattningssamling/skollag-2010800_sfs-2010-800.

19 Hallsén S. The Rise of Supplementary Education in Sweden: Arguments, Thought Styles, and Policy Enactment. ECNU Review of Education 2021; 4: 476–93.

20 European Union. Regions in the European Union Nomenclature of territorial units for statistics (NUTS) 2024 edition. Publications Office of the European Union 2024. DOI:10.2785/714519.

21 Library WO, Gasparrini A. Modeling exposure–lag–response associations with distributed lag non-linear models. Stat Med 2014; 33: 900–900.

22 Gasparrini A, Armstrong B, Kenward MG. Distributed lag non-linear models. Stat Med 2010; 29: 2224– 34.

23 Janes H, Sheppard L, Lumley T. Case-crossover analyses of air pollution exposure data: Referent selection strategies and their implications for bias. Epidemiology 2005; 16: 717–26.

24 Armstrong BG, Gasparrini A, Tobias A. Conditional Poisson models: a flexible alternative to conditional logistic case cross-over analysis. BMC Medical Research Methodology 2014 14:1 2014; 14: 122-.

25 Ni W, Agewall S, Benson L, et al. Cold Spells, Heat Waves, and Nonoptimal Air Temperature and Risk of Heart Failure Hospitalization in Sweden. JACC 2026; 88: 1016–29.

26 Guo Y, Gasparrini A, Armstrong B, et al. Global variation in the effects of ambient temperature on mortality: a systematic evaluation. Epidemiology 2014; 25: 781.

27 Ballester J, Quijal-Zamorano M, Méndez Turrubiates RF, et al. Heat-related mortality in Europe during the summer of 2022. Nat Med 2023; 29: 1857–66.

28 Xu Z, Huang C, Turner LR, Su H, Qiao Z, Tong S. Is Diurnal Temperature Range a Risk Factor for Childhood Diarrhea? PLoS One 2013; 8: e64713.

29 Hu Y, Cheng J, Yin Y, et al. Association of childhood asthma with intra-day and inter-day temperature variability in Shanghai, China. Environ Res 2022; 204. DOI:10.1016/j.envres.2021.112350.

30 Zhang Z, Xu D, Chen J, Meng Q, Liang Z, Zhang X. Daily diurnal temperature range associated with outpatient visits of acute lower respiratory infection in children: A time-series study in Guangzhou, China. Front Public Health 2022; 10: 951590.

31 Gronlund CJ. Racial and socioeconomic disparities in heat-related health effects and their mechanisms: a review. Curr Epidemiol Rep 2014; 1: 165.

32 Benmarhnia T, Deguen S, Kaufman JS, Smargiassi A. Vulnerability to Heat-related Mortality: A Systematic Review, Meta-analysis, and Meta-regression Analysis. Epidemiology 2015; 26: 781–93.

33 Vopham T, Hart JE, Laden F, Chiang YY. Emerging trends in geospatial artificial intelligence (geoAI): Potential applications for environmental epidemiology. Environ Health 2018; 17: 1–6.

34 Agarwal R, Melnick L, Frosst N, et al. Neural Additive Models: Interpretable Machine Learning with Neural Nets. Adv Neural Inf Process Syst 2020; 6: 4699–711.

35 Folkhälsomyndigheten. Impact on health of climate change in Sweden - A risk and vulnerability analysis. 2024 https://www.folkhalsomyndigheten.se/contentassets/477b2a19f4ae49c9a62e23078e6319dc/impact-on-health-of-climate-change-in-sweden.pdf (accessed Sept 23, 2025).

