## supplementary for "Climate factors and child health in Sweden: a nationwide whole-population study of 4.6 million school-aged children"

### Supplementary methods

### Description of Cohort

We conducted a whole-population case time-series study of individuals contributing follow-up time from the start of the calendar year in which they turned six years (or from first registration in Sweden after age six, if later) until the end of the calendar year in which they turned 18 years, death, emigration/deregistration, or the end of the study period on 31 December 2018, whichever came first. Health outcomes were obtained from National Patient Register (NPR) data, restricted to inpatient cases and to the years 1987–2018. The study cohort was drawn from all individuals born in Sweden between 1969 and 2018, totalling 6,722,066 persons identified from the national registers, of whom 4,638,247 individuals contributed follow-up time during the period of interest (ages 6 to 18 years). A total of 2,029,276 hospitalisation events were initially identified, occurring among 1,071,069 unique individuals. After excluding events not meeting the 28-day washout criteria, 1,737,334 events were retained for analysis. Following additional exclusion of events for which registered geographical information was not available for the event year, for example, where early or late registration in Sweden meant an individual's parish or district was not yet captured by LISA or RTB for that year 1,736,169 events remained, corresponding to 1,070,702 individuals.

The population is stratified into three school-age groups reflecting the Swedish education system stages to capture developmental and curricular differences. The early group (ages 6–9, roughly four years) covers the compulsory preschool class and the first years of compulsory schooling (Förskoleklass and Years 1–3). The middle group (ages 10–13, about four years) corresponds to the intermediate compulsory years (Years 4–7). The late group (ages 14–18, approximately five years) includes the final compulsory years and all upper secondary education (Years 8–9 and Gymnasium Years 10–12), following the structure defined in the Swedish Education Act.^1^

Calendar period was categorised a priori into three intervals (1987–1993, 1994–2005, 2006–2018) of broadly similar length, the first two separated by the major Swedish education reforms of the early 1990s.^2^ The periods are used to assess whether associations changed over time and are not intended to represent distinct welfare-policy regimes.

Table S1: Study period based on socioeconomic and policy phases influencing social determinants of education in Sweden

| **Period** | **Key Features** | **Relevant Policy and Economic Events** |
| --- | --- | --- |
| 1987-1993 | Economic Crisis and Structural Reform | Severe financial crisis, deregulation, austerity measures challenging the welfare state |
| 1994-2005 | Market-Oriented Reforms and Deregulation | Expansion of school choice, introduction of market principles, increased socioeconomic stratification |
| 2006-2018 | Focus on Educational Equity | Policies targeting improved equity, supplementary education, and support for underperforming students. |

Cause-specific outcomes were constructed by grouping diagnosis codes into harmonised disease categories using the chapter-level classifications of the International Classification of Diseases, Ninth and Tenth Revisions (ICD-9 and ICD-10). Corresponding ICD-9 and ICD-10 code ranges were mapped to a common set of 22 cause categories (Table S2). The four most frequent categories among children were injury and poisoning, symptoms and ill-defined conditions, respiratory diseases, and digestive diseases. Of these, injury and poisoning, respiratory diseases, and digestive diseases were analysed as secondary outcomes in the main text, alongside mental disorders (the fifth most frequent category); symptoms and ill-defined conditions was excluded as a non-specific residual chapter not amenable to clinical interpretation (see Methods).

Parental medical history was defined as any ICD-coded diagnosis recorded for either parent in the National Patient Register before the child's birth, coded as a binary indicator (yes/no). Codes were included where the underlying condition could plausibly have had a severe impact on the parent's own health, or on the health or development of the child; for example, genetic disorders and psychiatric conditions. Because this definition captures a broad range of conditions, the variable should be interpreted as a general indicator of significant parental morbidity rather than as a diagnosis-specific exposure.

Geographically stratified analyses used the NUTS1 European Union classification of Swedish regions, known locally as *landsdelar*, based on each individual's registered residence, providing a broad regional framework for examining spatial differences in climate exposures and health outcomes.^3^

Admission patterns during school holidays may differ from term time for reasons unrelated to climate (for example, changes in daily activity, travel and healthcare-seeking behaviour). To assess whether these periods influenced our estimates, we repeated the analysis separately for holiday and non-holiday periods. Holiday periods were defined as the Swedish summer break, from Midsummer to the end of August (19 June – 31 August), and the Christmas break (15 December to 9 January). Because each stratum is a calendar month, holiday status was assigned to the stratum according to the date of the index admission, not day by day. This keeps the classification constant within each stratum, as required by the conditional Poisson model. As a result, a stratum whose admission fell on or after 19 June was classified as a holiday stratum in full, including days of that month before the break began. In total, 382,113 of 1,736,169 admissions (22·0%) fell in holiday periods and 1,354,056 (78·0%) in non-holiday periods.

### Population Weighted Mean Climate Data

There have been regular changes between parish boundaries every year in Sweden. 1 January 2000, the Church of Sweden was separated from the state, and since then the Church of Sweden has decided on the division of parishes^4^. Parish boundaries in Sweden have changed at various points during the study period. On 1 January 2000, the Church of Sweden was separated from the state, after which the Church assumed responsibility for parish boundary decisions.^4^ Before 2000, the parish division was stable; between 2000 and 2015, however, many parishes were merged into larger units, and the total number of parishes decreased by approximately 2,515 to around 1,365.^5^ From 2015, Sweden shifted from parishes to districts as the official administrative boundary unit.^6^ In this study, all historical parish codes, and district codes introduced from 2015 onward, were harmonised to the 2015 parish-code reference framework to standardise administrative boundaries across the study period. Using this reference level, daily climatic exposures were estimated at the parish level via calculations of population-weighted mean variables.

$$\bar{x}_{j,t} = \frac{\sum_{i\in I_{j}} ({POP}_{i}\cdot A_{ij}) x_{i,t}}{\sum_{i\in I_{j}} ({POP}_{i}\cdot A_{ij})}$$

Where, $\bar{x}_{j,t}$ is population-weighted mean exposure in administrative unit $j$on day $t$. $x_{j,t}$ extracted (polygon-mean) climate value for intersected polygon $i$on day $t$. $I_{j}$set of all intersected polygons $i$that fall within administrative unit $j$. ${POP}_{i}$ population attribute for the population-grid polygon $i$(as used in weighting). $A_{ij}$ area of overlap (intersection) between population polygon $i$and administrative unit $j$, in $\text{km}^{2}$(i.e., $A_{ij} = {area}_{ij}/{10}^{6}$if area is computed in $\text{m}^{2}$).

Population distribution data were linked based on polygons of reference parishes, and overlay analyses were conducted to derive weighted daily mean temperatures for each parish based on the 2015 boundaries ^7^. These daily exposure values were subsequently aggregated to the individual level for analysis. Daily ambient temperature and relative humidity were used as extracted from SMHIGridClim without further unit conversion.

To quantify intraday temperature variability, we used sub-daily air temperatures at 6-hour intervals (00:00, 06:00, 12:00, and 18:00 local time). For each day $d$, with temperatures$T_{d-1,18}$, $T_{d,00}$, $T_{d,06}$, $T_{d,12}$, and $T_{d,18}$, we defined intraday variability as the sum of absolute successive 6-hour changes:

$${ITV}_{d}=\mid T_{d,00}-T_{d-1,18}\mid+\mid T_{d,06}-T_{d,00}\mid+\mid T_{d,12}-T_{d,06}\mid+\mid T_{d,18}-T_{d,12}\mid$$

Higher values indicate greater within-day temperature fluctuations; the unit is °C/day.

To characterise within-day temperature variability, we compared two commonly used metrics: the diurnal temperature range (DTR) and ITV. DTR was defined as the difference between the daily maximum and minimum temperature $(T_{max}- T_{min})$, capturing the daily thermal amplitude based only on two extrema. Although DTR and ITV were correlated, time-series comparisons illustrated that ITV can be elevated on days with multiple rapid swings even when the daily max–min range is modest (Supplementary Figure S8), whereas DTR does not capture such intra-day reversals. In seasonal DLNM analyses (Supplementary Figure S7), ITV produced more clearly non-linear exposure–response functions than DTR and provided a more direct representation of frequent within-day temperature changes; therefore, ITV was retained as the primary metric of temperature variability in the main analyses.

Daily precipitation (pr) in SMHIGridClim is provided as a flux ($kgm^{-2}s^{-1}$). We converted this to daily accumulated precipitation (mm/day) using the water-equivalent relationship (1 ${kgm}^{-2}s^{-1}$ = 1 mm) and multiplying by seconds per day:

$$P_{d}\text{ }(mm/day)=pr_{d}\text{ }(\mathrm{kg}\text{ }m^{-2}\text{ }s^{-1})\times86400$$

Although SMHI defines heatwaves in Sweden when daily max temperature crosses 25 °C for three consecutive days but general definition of climate extremes for all variables are not clearly defined. Hence for this study, Climate extremes have been defined as values below 1^st^ and above 99^th^ percentiles for Generalisability.

### Data Sources

*Geographic Data:* Annual parish and district codes for geographical linkage were extracted from administrative population data compiled by SCB in LISA register, allowing the assignment of spatial location to individuals over time. Vector boundaries for parishes (församlingar) and districts (distrikt) were acquired from Lantmäteriet, the Swedish mapping, cadastral, and land registration authority. These shapefiles include detailed spatial delineations and metadata documenting the transition and reconciliation of district boundaries to the consistent 2015 parish-level administrative units, ensuring harmonized spatial referencing across temporal changes in administrative geography.

*Climate Data:* Climate exposure data were derived from the SMHI Gridded Climatology dataset (SMHIGridClim) covering the years 1961 to 2018. This dataset spans the Nordic countries including Sweden, Finland, and Norway on a 2·5 km by 2·5 km horizontal grid.^8^ Air temperature is at 1-, 3-, or 6-hour temporal resolutions depending on the time period, as well as daily minimum and maximum temperatures and daily precipitation totals relevant to this study.

### Time Series Data

Administrative codes corresponding to each event year were assigned to the data. When codes were missing for certain years from 2015’s parish codes, the reference table of code changes which was previously used in creating the climate exposure dataset, was applied for standardization. The observation window for each event was expanded to include the entire month of the event date plus a 21-day lag period. Incorporating a lag period acknowledges that the health impact of temperature may not be immediate but can manifest after several days due to biological and physiological response times.^9^ The outcome variable was coded as 1 on the event date and 0 for all other days within the expanded window. Subsequently, temperature exposure data and covariates required for stratification were linked to the dataset based on administrative codes and dates. The final time series dataset comprised the following variables: individual identifier (lopnr), outcome status, date, sex, income quintile, parental education levels (both father and mother), age group, and calendar period.

### Characteristics of Outcome

Among 1,070,702 children with at least one inpatient admission, 2,029,276 hospitalisation events were initially identified; after enforcing the 28-day washout, 1,737,334 events remained, and a further 1,736,169 events remained after excluding those with no location information (Table S1). Of these retained events, 51·9% occurred in males (901,157) and 48.1% in females (835,012), and events were most frequent among adolescents aged 14–18 years (44·6%, 774,423), followed by ages 6–9 (28·5%, 494,520) and 10–13 (26·9%, 467,226). Using the specified ICD groupings, injury and poisoning accounted for 23.7% of events (411,024), respiratory diseases for 11·.1% (191,997), digestive diseases for 8·6% (149,150), and mental disorders for 6·2% (107,890), while the remaining 50·5% (876,108) fell outside these categories; the largest share of these (233,304 events, 13·4% of all retained events) were classified as symptoms and ill-defined conditions, a non-specific residual category not analysed separately. When categorised by outcome type, acute events represented 5·54% (96,244) and chronic events 2·64% (45,854), with 91·81% classified as none (1,594,071). Event distributions by sociodemographic factors were similar to the underlying hospitalised population: 50·9% of events occurred among children with parental upper secondary education, income quintiles were approximately evenly distributed, and the largest regional contribution was from South Sweden (46·3%). The largest share of events occurred during 1994–2005 (40.8%), and 55·5% of events were among children with parental medical history = no (42·7% yes; 1·8% missing).

For clarity in figures, we truncated the exposure–response curves to the plotting ranges of climatic variables specified in Table S2.

**Case time-series design**

Analyses were restricted to individuals who experienced the outcome. Children could be admitted more than once during follow-up. Because admissions occurring close together often represent readmission or continuation of the same episode rather than a new, independent event, we applied a 28-day washout: any hospitalisation within 28 days of a previous one was excluded, and only admissions separated by at least 28 days were retained as distinct events. Each retained event formed its own stratum, comprising the days of the same individual within the same calendar month and year. Comparisons were therefore made only between days within the same individual and calendar month, which accounts for within-person clustering and controls for all confounders that are constant within that stratum. Stratum-specific intercepts were eliminated by conditioning in a fixed-effect Poisson model.

**Rationale.** The case time-series design^10^ extends self-matched case-only methods, such as the case-crossover and self-controlled case series, to longitudinal data. Data are arranged as a set of individual time series, one for each case, and analysed with the regression methods used in conventional time-series analysis. This retains the main strength of self-matched designs, which is control of time-invariant confounding by design, together with the flexibility of time-series models to represent non-linear and lagged exposure–response associations through distributed lag non-linear models.^10,11^ Unlike the time-stratified case-crossover design, which compares an event day with a small set of pre-selected referent days, the case time-series uses every day in the stratum. It also allows time-varying covariates to be included directly.^12–14^

**Event definition and washout.** Analyses were restricted to individuals who experienced the outcome. Children could be admitted more than once during follow-up. Admissions occurring close together often represent readmission or continuation of the same episode rather than a new, independent event. We therefore applied a 28-day washout: any hospitalisation within 28 days of a previous one was excluded, and only admissions separated by at least 28 days were retained as distinct events.

**Stratification.** Each retained event defined its own stratum, comprising all days of the same individual within the same calendar month and year. Each stratum therefore contained exactly one event day and 27 to 30 non-event days. Comparisons were made only between days within a stratum. This controls for all characteristics of the child that are constant within the month (for example sex, age group, parental education and income, underlying health, and residence) and for slowly varying factors such as long-term trends and broad seasonality. Because each event forms its own stratum, repeated events in the same child are analysed as separate comparisons, and within-person clustering is accounted for by design.

**Model.** Daily outcome indicators were modelled with a fixed-effect (conditional) Poisson regression, in which stratum-specific intercepts were eliminated by conditioning rather than estimated^12^. Conditioning on the stratum total gives estimates equivalent to those from conditional logistic regression, but it is computationally feasible at the scale of this study (1.7 million strata) and accommodates the distributed lag cross-basis directly. Residual within-month variation was controlled with indicator terms for day of week and week of year (see main text). Effect modifiers that are constant within a stratum cannot enter the model as main effects. Their modifying role was therefore assessed through interactions between the cross-basis and each modifier (Supplementary Table S4).

**Assumptions.** The design rests on four assumptions: (i) time-varying confounding within each stratum is adequately controlled by the day-of-week and week-of-year terms; (ii) the occurrence of an event does not alter the probability of a subsequent event within the retained series, an assumption supported by the 28-day washout and assessed in a sensitivity analysis restricted to each child's first hospitalisation; (iii) exposure measurement error is non-differential with respect to event timing; and (iv) each child's registered parish approximates their location on the exposure days, so that assigned climate values approximate true exposure.

**Relation to the sensitivity analysis.** The time-stratified case-crossover analysis (Supplementary Figure S11) uses the same individual-month strata but restricts comparison days to those falling on the same weekday as the event23,24. It therefore provides a check on whether the results depend on using all days in the stratum.

Inclusion-Exclusion Criteria

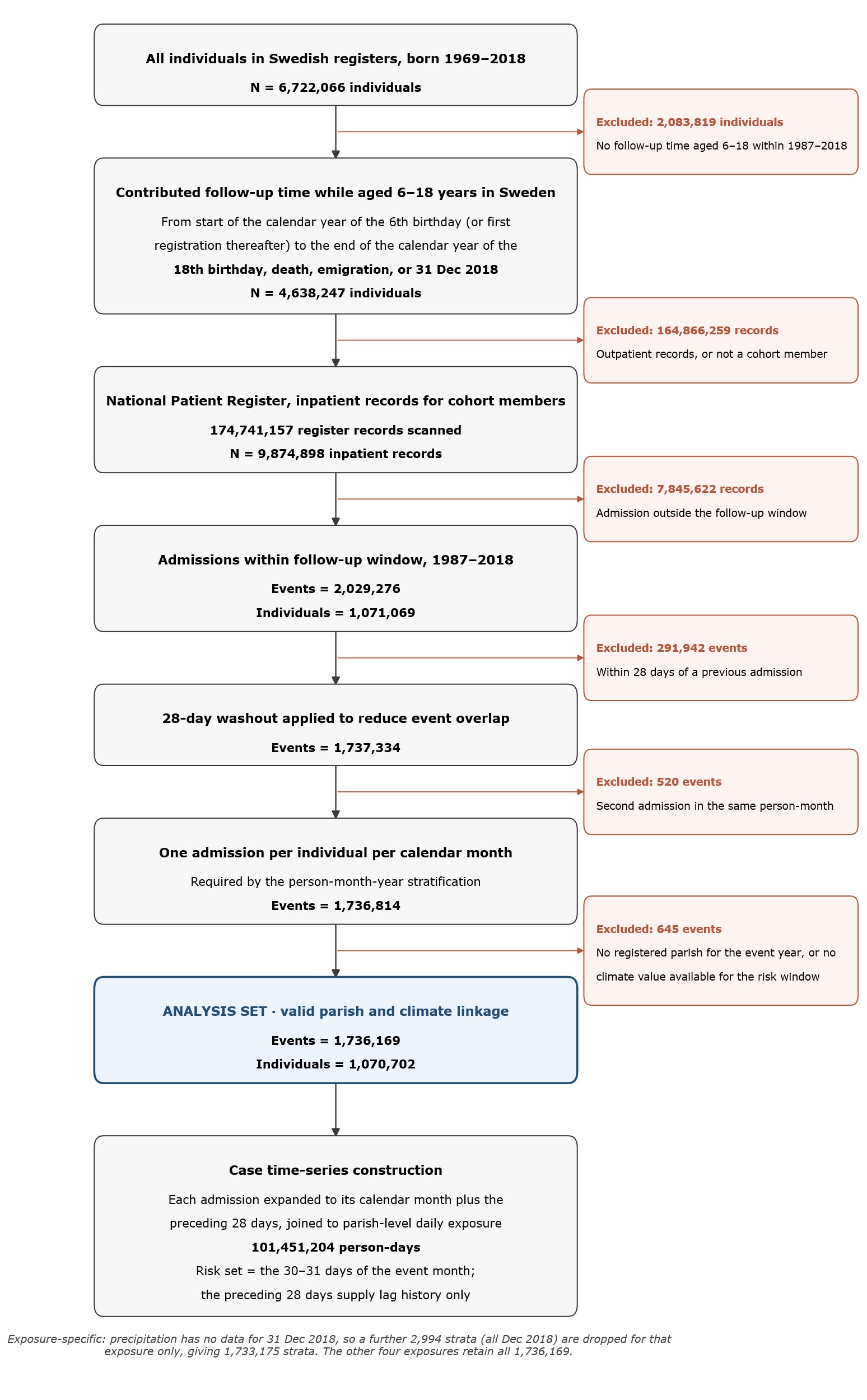

Table S2: Characteristics of outcomes

| **Characteristic** | **Category** | **1,736,169 (N%)** | **Outcomes dropped for Precipitation** |
| --- | --- | --- | --- |
| ***Sex*** | | |  |
|  | Female | 835,012 (48.10%) | 1,504 |
|  | Male | 901,157 (51.90%) | 1,490 |
| ***Age group (years)*** | | |  |
|  | 06-09 | 494,520 (28.48%) | 743 |
|  | 10-13 | 467,226 (26.91%) | 698 |
|  | 14-18 | 774,423 (44.61%) | 1,553 |
| ***Parent's education*** | | |  |
|  | Primary school | 207,106 (11.93%) | 316 |
|  | Upper secondary | 884,236 (50.93%) | 1,254 |
|  | University | 606,076 (34.91%) | 1,331 |
|  | Missing | 38,751 (2.23%) | 93 |
| ***Income quintile*** | | |  |
|  | 1 (Lowest) | 301,164 (17.35%) | 547 |
|  | 2 | 335,914 (19.35%) | 626 |
|  | 3 | 350,514 (20.19%) | 654 |
|  | 4 | 355,746 (20.49%) | 566 |
|  | 5 (Highest) | 357,540 (20.59%) | 534 |
|  | Missing | 35,291 (2.03%) | 67 |
| ***Regions (NUTS1)*** | | |  |
|  | East Sweden | 567,364 (32.68%) | 1,055 |
|  | South Sweden | 803,075 (46.26%) | 1,345 |
|  | North Sweden | 365,730 (21.07%) | 594 |
| ***Time period*** | | |  |
|  | 1987-1993 | 396,263 (22.82%) | ·· |
|  | 1994-2005 | 709,025 (40.84%) | ·· |
|  | 2006-2018 | 630,881 (36.34%) | 2,994 |
| ***Outcome type*** | | |  |
|  | Acute | 96,244 (5.54%) | 192 |
|  | Chronic | 45,854 (2.64%) | 62 |
|  | None | 1,594,071 (91.82%) | 2,740 |
| ***Parental Medical History*** | | |  |
|  | Yes | 740,409 (42.65%) | 1,449 |
|  | No | 963,805 (55.51%) | 1,480 |
|  | Missing | 31,955 (1.84%) | 65 |

Table S3: Harmonised cause-specific outcome categories based on corresponding ICD-9 and ICD-10 code ranges, with category descriptions and the number of records among children.

| **Category** | **ICD-9 code** | **ICD-10 code** | **Description** | **Records** |
| --- | --- | --- | --- | --- |
| Injury and poisoning | 800–999 | S00–T98 | Injury, poisoning and certain other consequences of external causes | 411,024 |
| Symptoms and ill-defined conditions | 780–799 | R00–R99 | Symptoms, signs and abnormal clinical and laboratory findings | 233,304 |
| Respiratory diseases | 460–519 | J00–J99 | Diseases of the respiratory system | 191,997 |
| Digestive diseases | 520–579 | K00–K93 | Diseases of the digestive system | 149,150 |
| Mental disorders | 290–319 | F00–F99 | Mental and behavioural disorders | 107,890 |
| Congenital conditions | 740–759 | Q00–Q99 | Congenital malformations, deformities and chromosomal abnormalities | 84,208 |
| Endocrine and metabolic diseases | 240–279 | E00–E90 | Endocrine, nutritional and metabolic diseases | 77,447 |
| Genitourinary diseases | 580–629 | N00–N99 | Diseases of the genitourinary system | 66,756 |
| Nervous-system diseases | 320–359 | G00–G99 | Diseases of the nervous system | 61,611 |
| Musculoskeletal diseases | 710–739 | M00–M99 | Diseases of the musculoskeletal system and connective tissue | 60,653 |
| Infectious and parasitic diseases | 001–139 | A00–B99 | Certain infectious and parasitic diseases | 51,087 |
| Health-service factors | V codes | Z00–Z99 | Factors influencing health status and contact with health services | 43,669 |
| Neoplasms | 140–239 | C00–D48 | Neoplasms | 39,242 |
| Ear diseases | 380–389 | H60–H95 | Diseases of the ear and mastoid process | 28,685 |
| Skin diseases | 680–709 | L00–L99 | Diseases of the skin and subcutaneous tissue | 21,382 |
| Circulatory diseases | 390–459 | I00–I99 | Diseases of the circulatory system | 20,945 |
| Pregnancy and childbirth | 630–679 | O00–O99 | Pregnancy, childbirth and the puerperium | 20,357 |
| Blood and immune disorders | 280–289 | D50–D89 | Diseases of the blood and blood-forming organs and certain immune disorders | 18,175 |
| Eye disorders | 360–379 | H00–H59 | Disorders of the eye and adnexa | 16,087 |
| Perinatal conditions | 760–779 | P00–P96 | Certain conditions originating in the perinatal period | 1,197 |

### Supplementary Figures

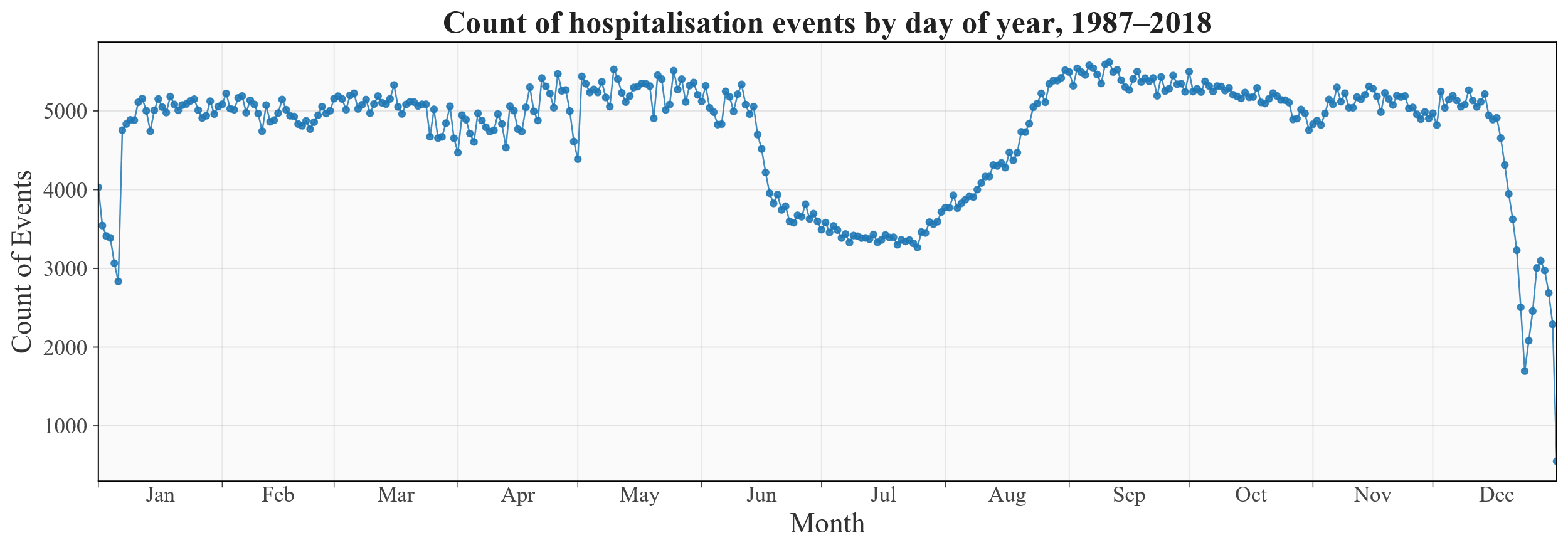
 *(a)*
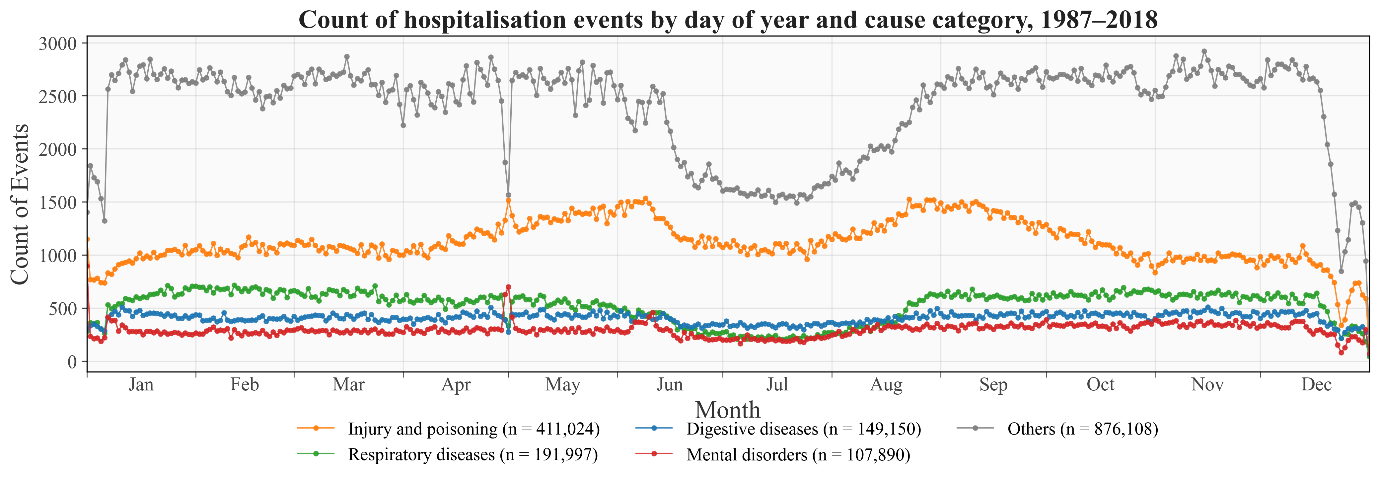
 *(b)*

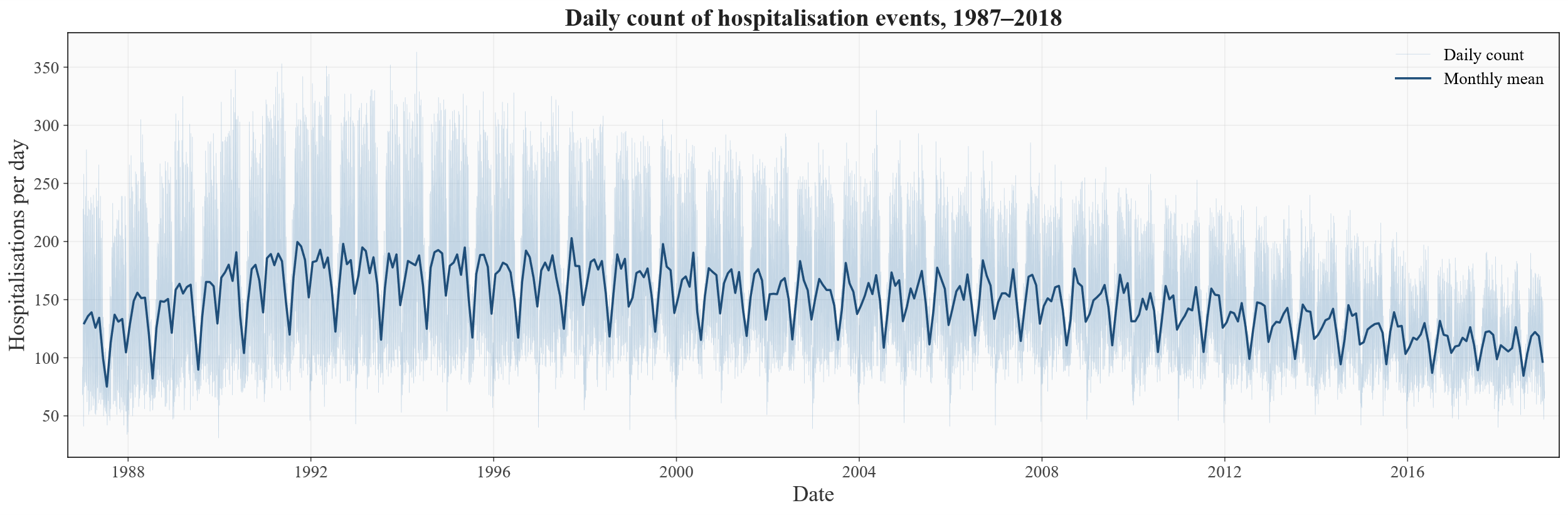
 *(c)*

Figure S1: **Distribution of hospitalisation events over time, 1987–2018.** (a) Total count of hospitalisation events by day of year, pooled across all study years. (b) Count of events by day of year and cause category, pooled across all study years: injury and poisoning (n = 411,024), respiratory diseases (n = 191,997), digestive diseases (n = 149,150), mental disorders (n = 107,890), and all remaining ICD chapters combined ("Others", n = 876,108). (c) Daily count of events over calendar time; the faint line shows the count on each day and the bold line the mean daily count within each calendar month. All panels are based on the 1,736,169 hospitalisation events retained for analysis after the 28-day washout and exclusion of events with missing geographic information. In panels (a) and (b), day 366 occurs only in the eight leap years within the study period, so its total is approximately one quarter of that for adjacent days.

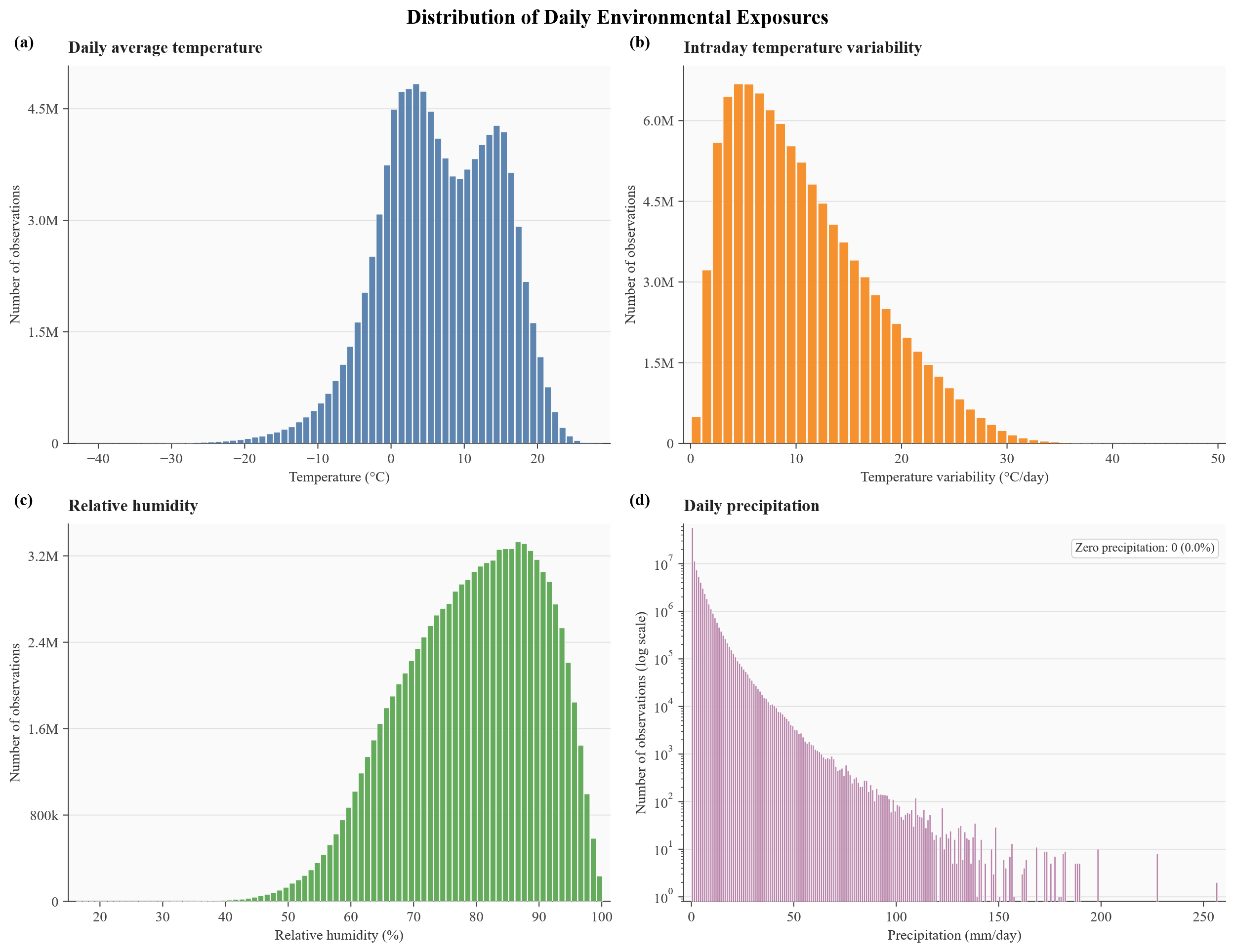

Figure S2: Histogram of exposure values at Lag = 0 for each event.

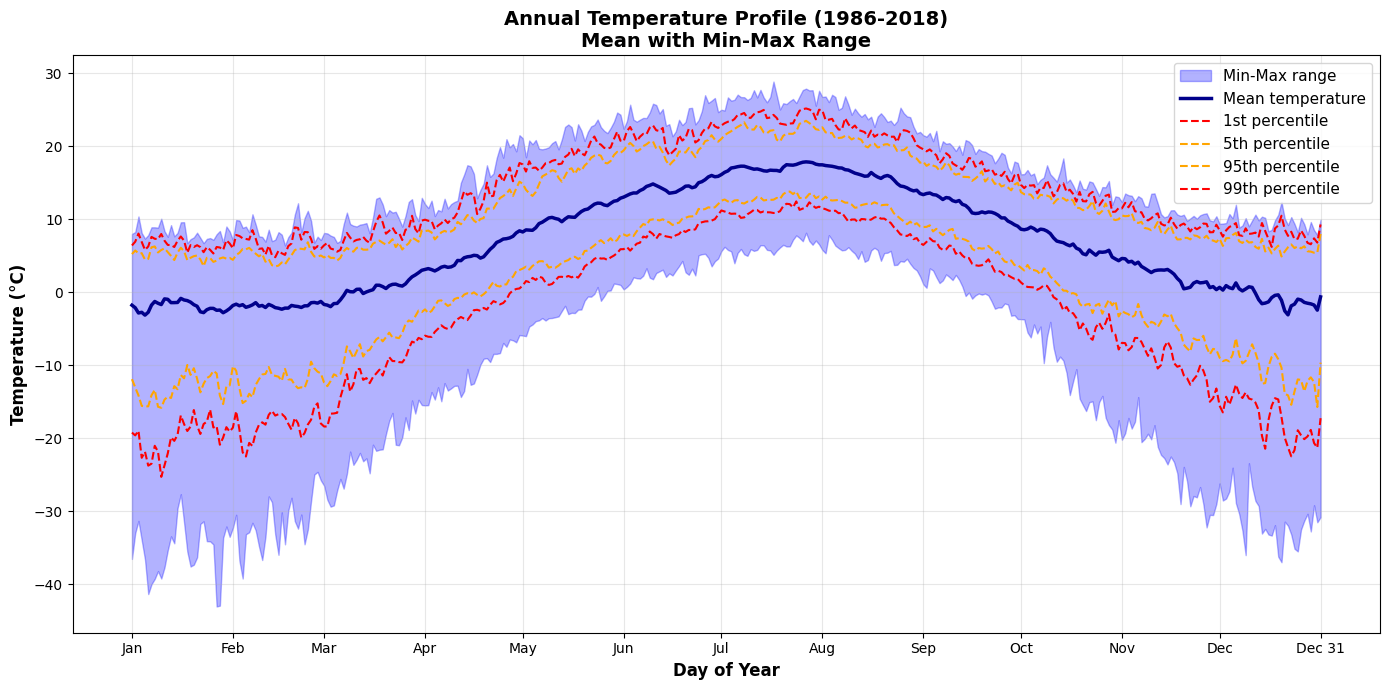

Figure S3: Annual Temperature Profile (1986-2018)

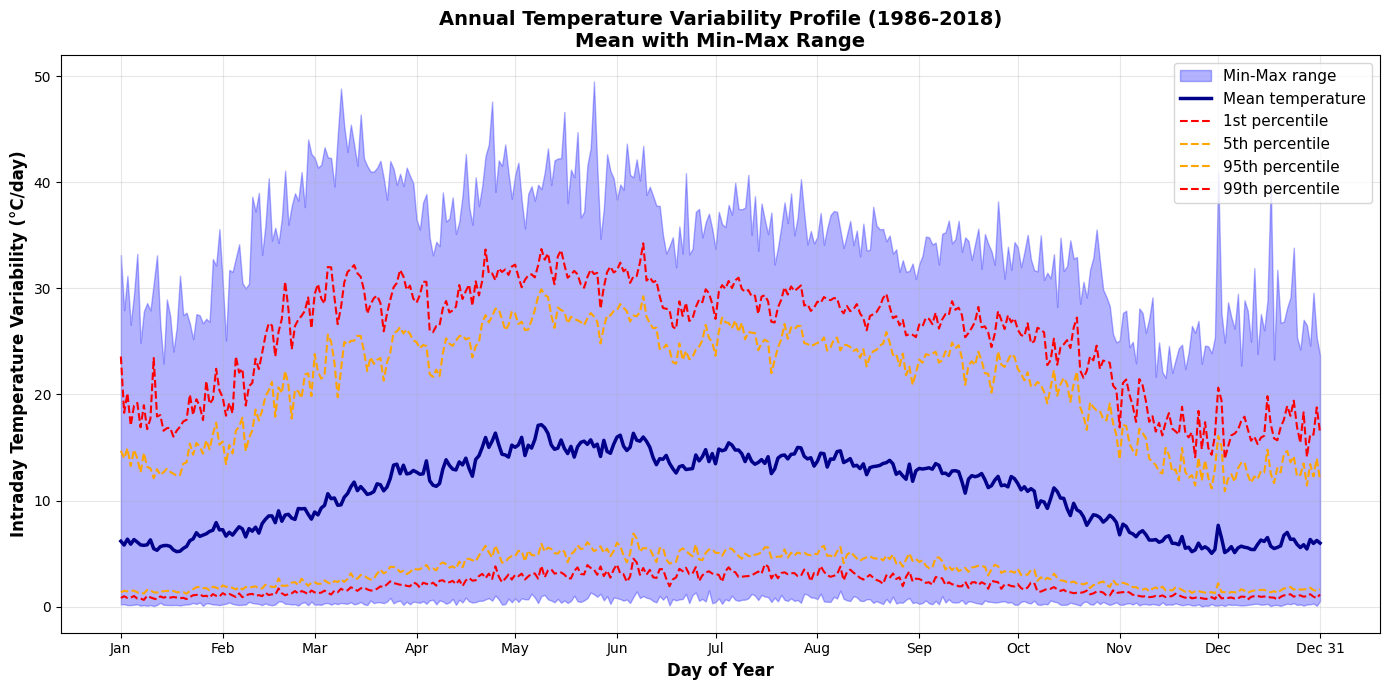

Figure S4: Annual Temperature Variability Profile (1986-2018)

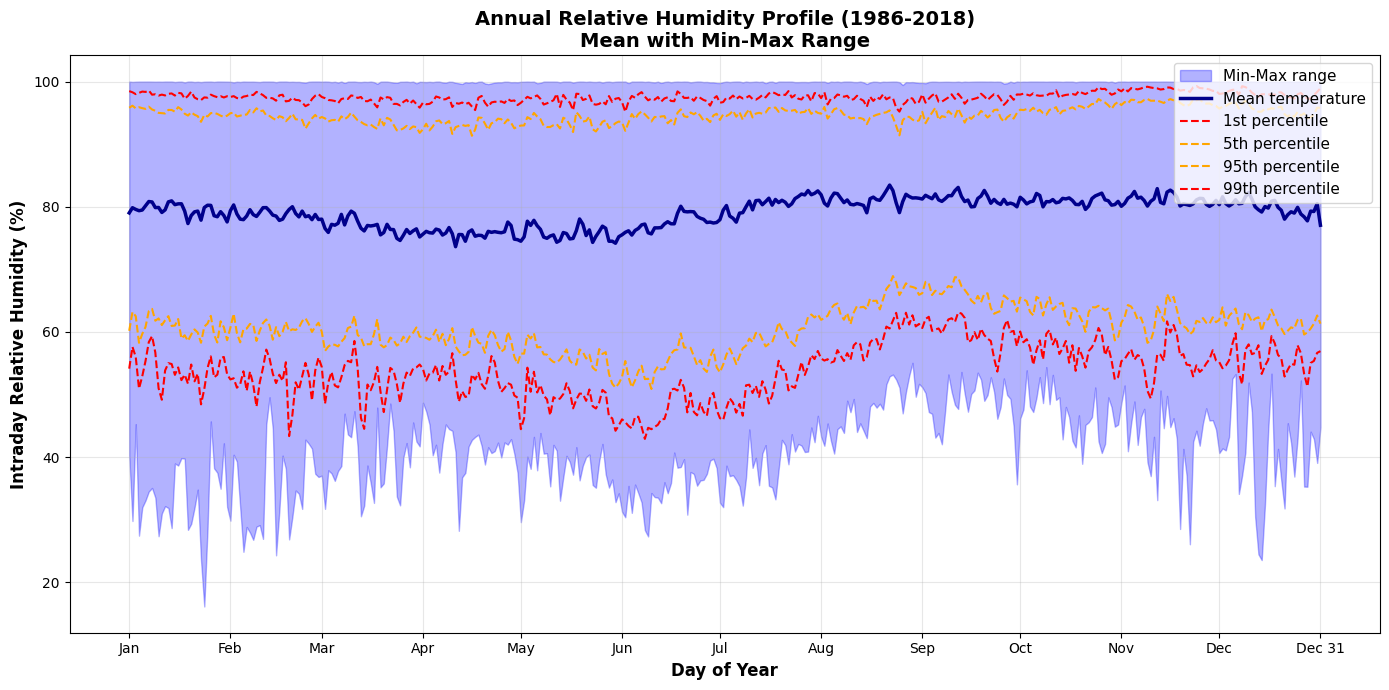

Figure S5: Annual Relative Humidity Profile (1986-2018)

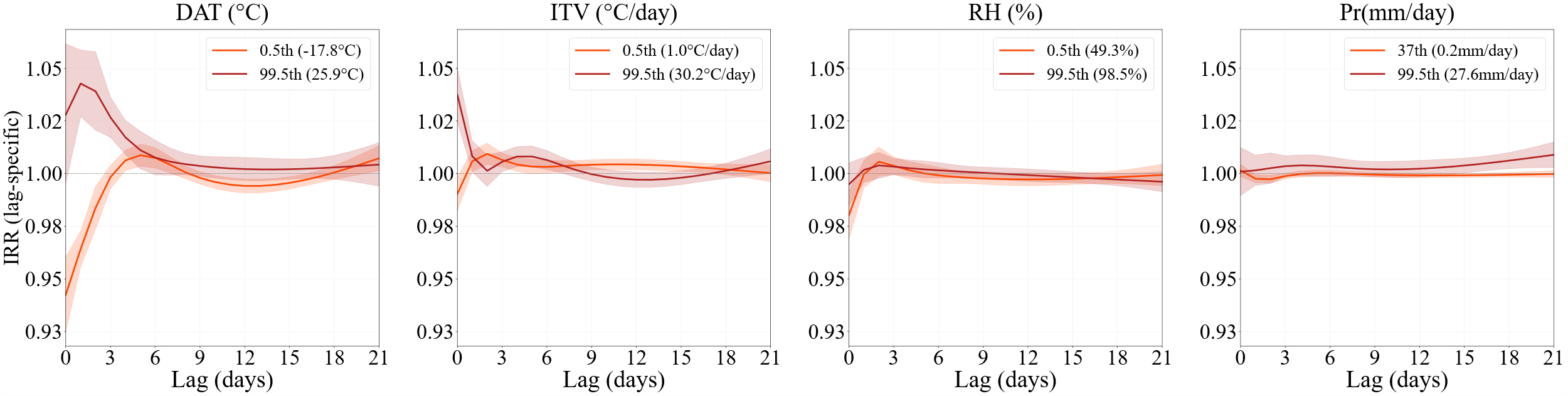

Figure S6: Lag-specific incidence rate ratios (IRR) for hospitalisation at low and high percentiles of each exposure, relative to the median, over lags 0–21 days: (a) daily average temperature, (b) intraday temperature variability, (c) relative humidity, (d) precipitation. The low percentile is the 1st, except for precipitation (37th). Shaded areas show 95% confidence intervals.

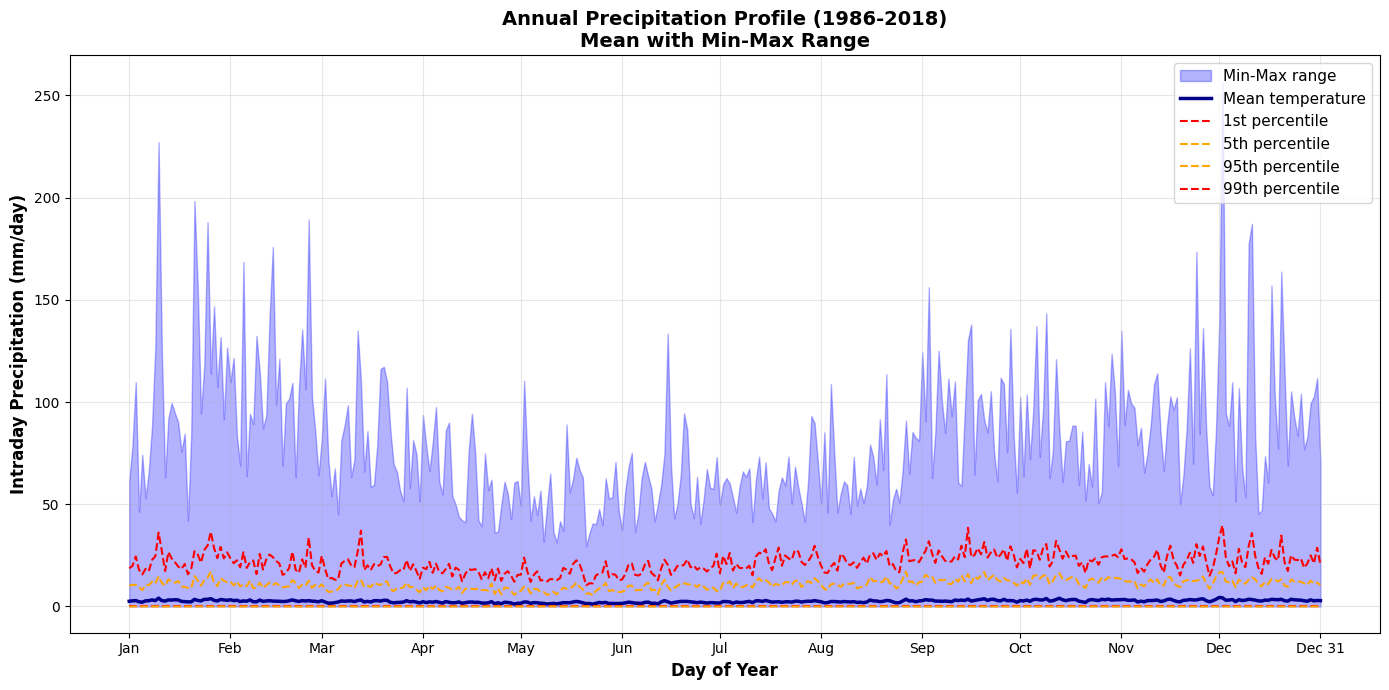

Figure S7: Annual Precipitation Profile (1986-2018)

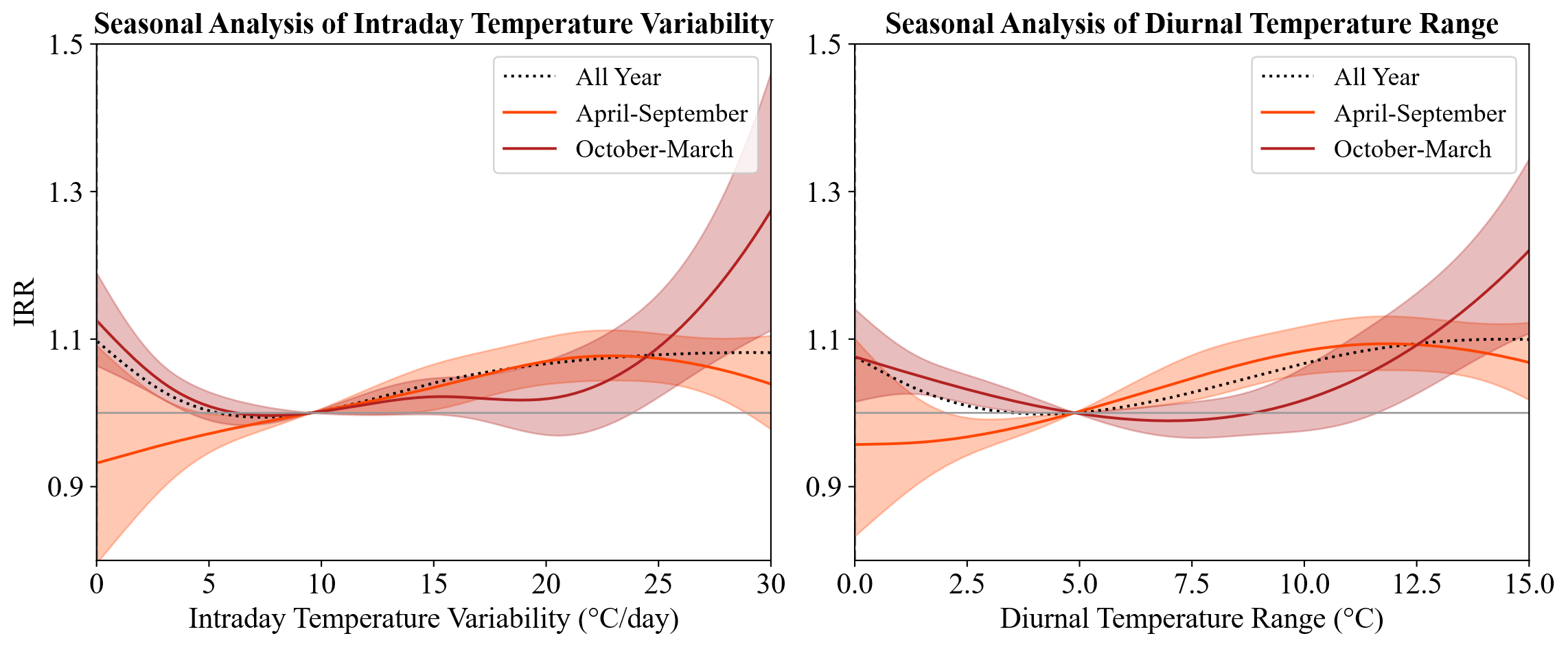

Figure S7: cumulative exposure-response associations for ITV (left) and DTR (right)

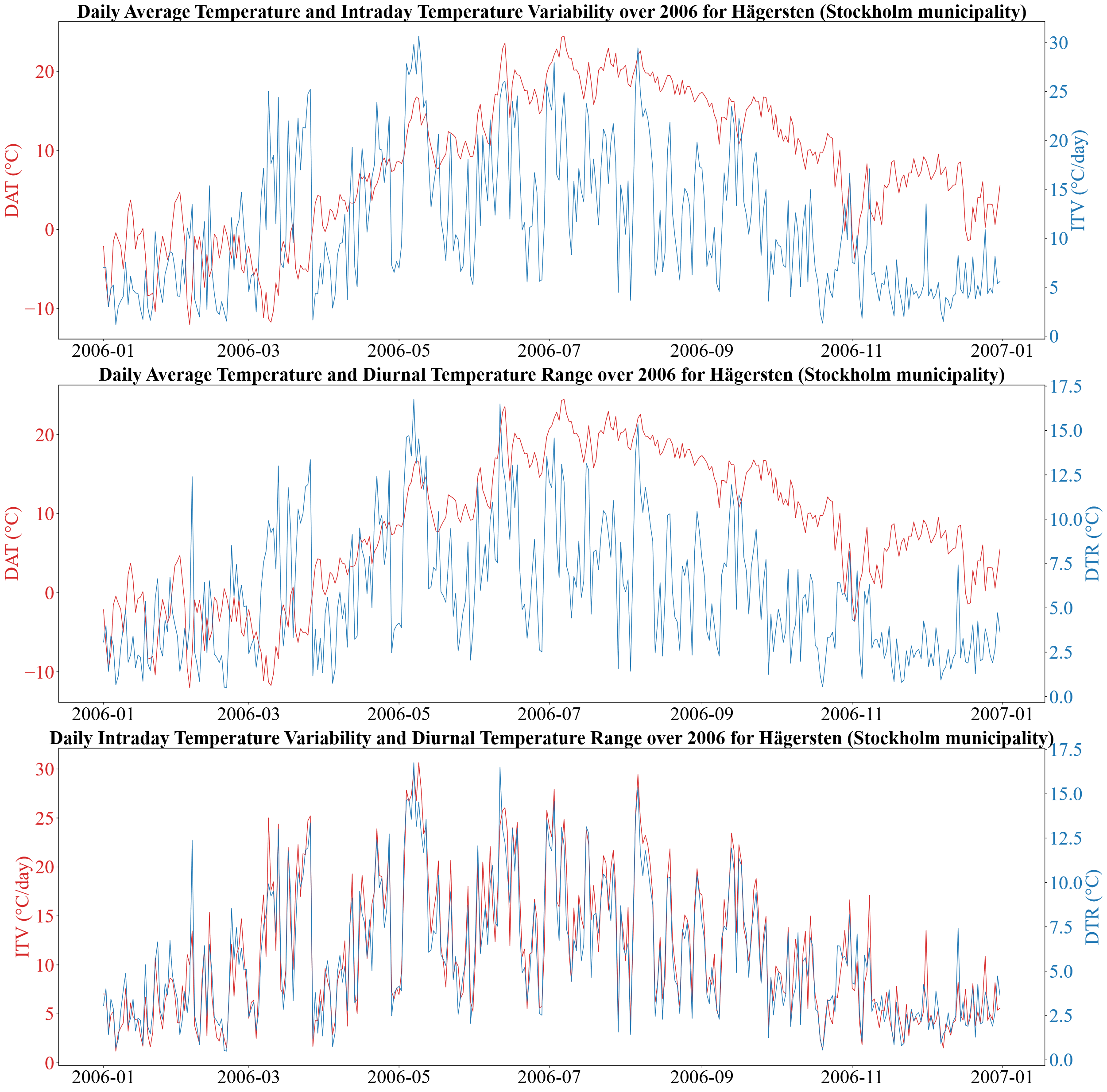

Figure S8: Comparison of season trend of Daily Average Temperature, Intraday Temperature Variability and Diurnal Temperature Range	over 2006 for Hägersten (Stockholm municipality)

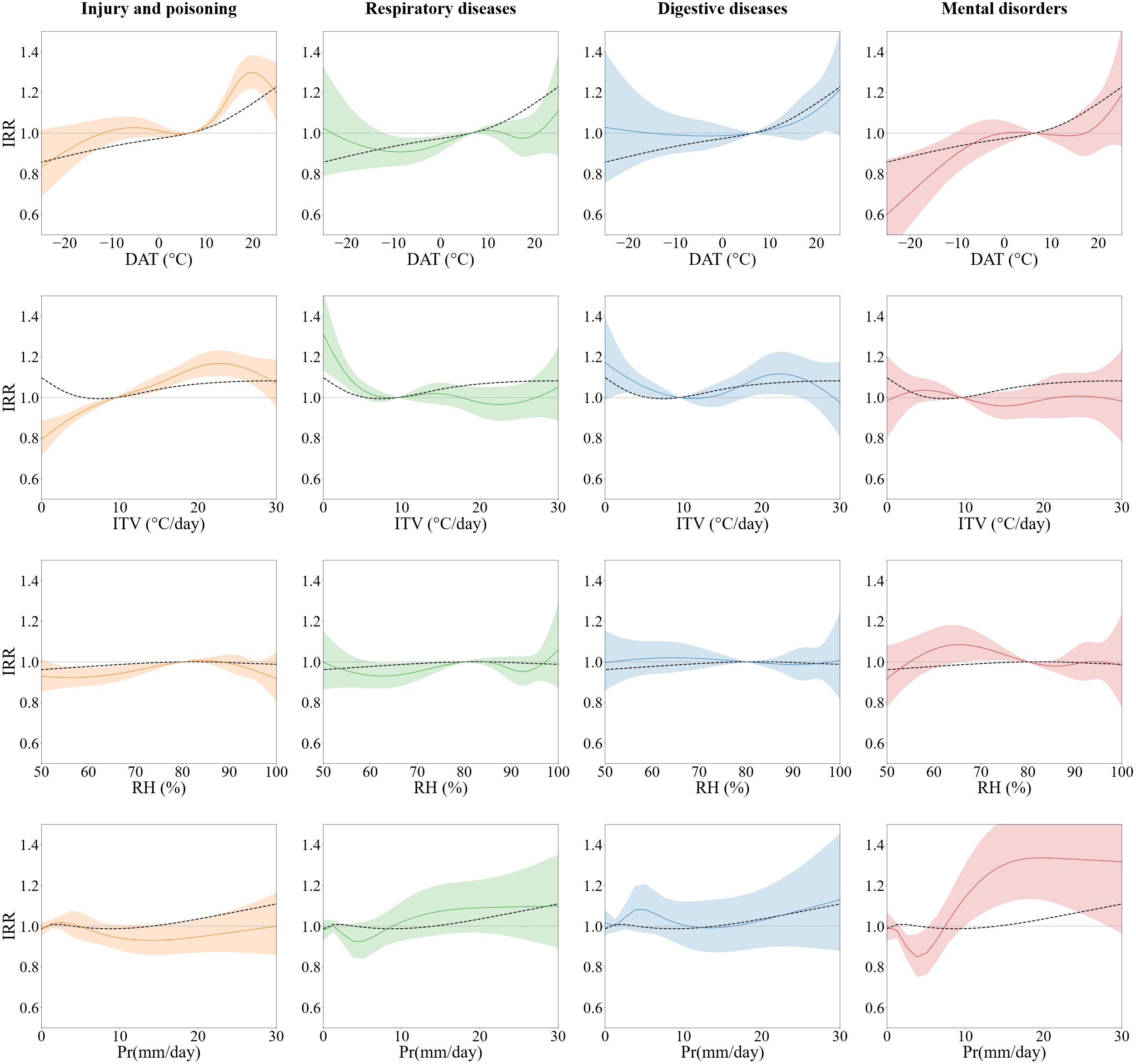

Figure S10: Outcome-specific 21-day cumulative exposure-response associations for different climate variables

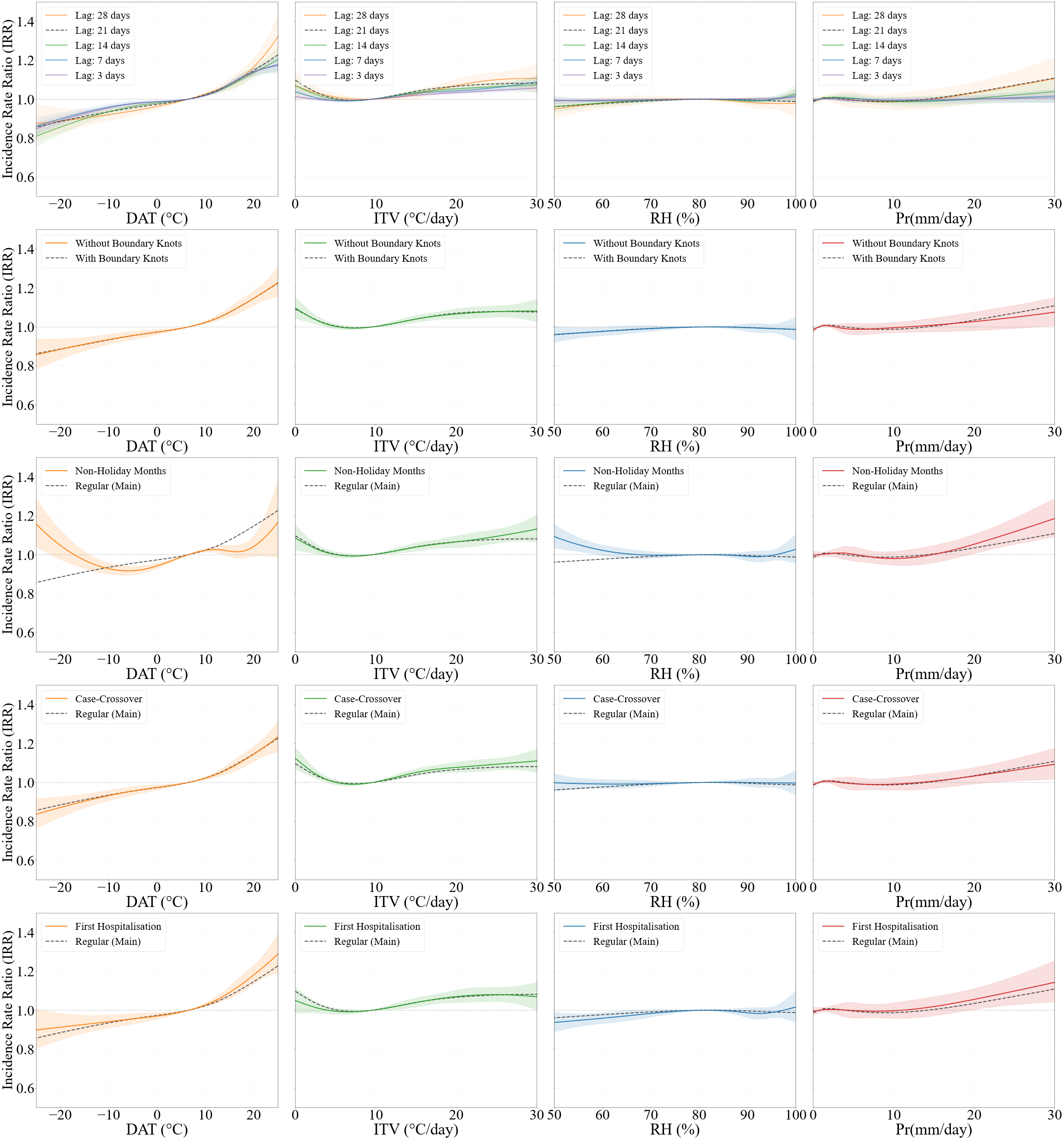

Figure S11: Sensitivity analyses for daily average temperature (DAT), intraday temperature variability (ITV), relative humidity (RH) and precipitation (columns, left to right). Rows, top to bottom: (i) alternative maximum lag periods of 3, 7, 14, 21 (main analysis) and 28 days; (ii) cross-basis specification with versus without boundary knots, placed at the 1st and 99th percentiles for DAT, ITV and RH and at the 37th and 99th percentiles for precipitation; (iii) restriction to non-holiday months, excluding the summer (19 June – 31 August) and Christmas (15 December – 9 January) school breaks; (iv) time-stratified case-crossover design, with control days on the same day of the week within the same calendar month; and (v) restriction to each child's first recorded hospitalisation during follow-up. In rows (ii)–(v), the coloured line shows the sensitivity analysis with its 95% confidence interval (shaded), and the black dashed line shows the main analysis. In row (i), the black dashed line is the 21-day main analysis. Estimates are cumulative incidence rate ratios (IRR) over the lag period.

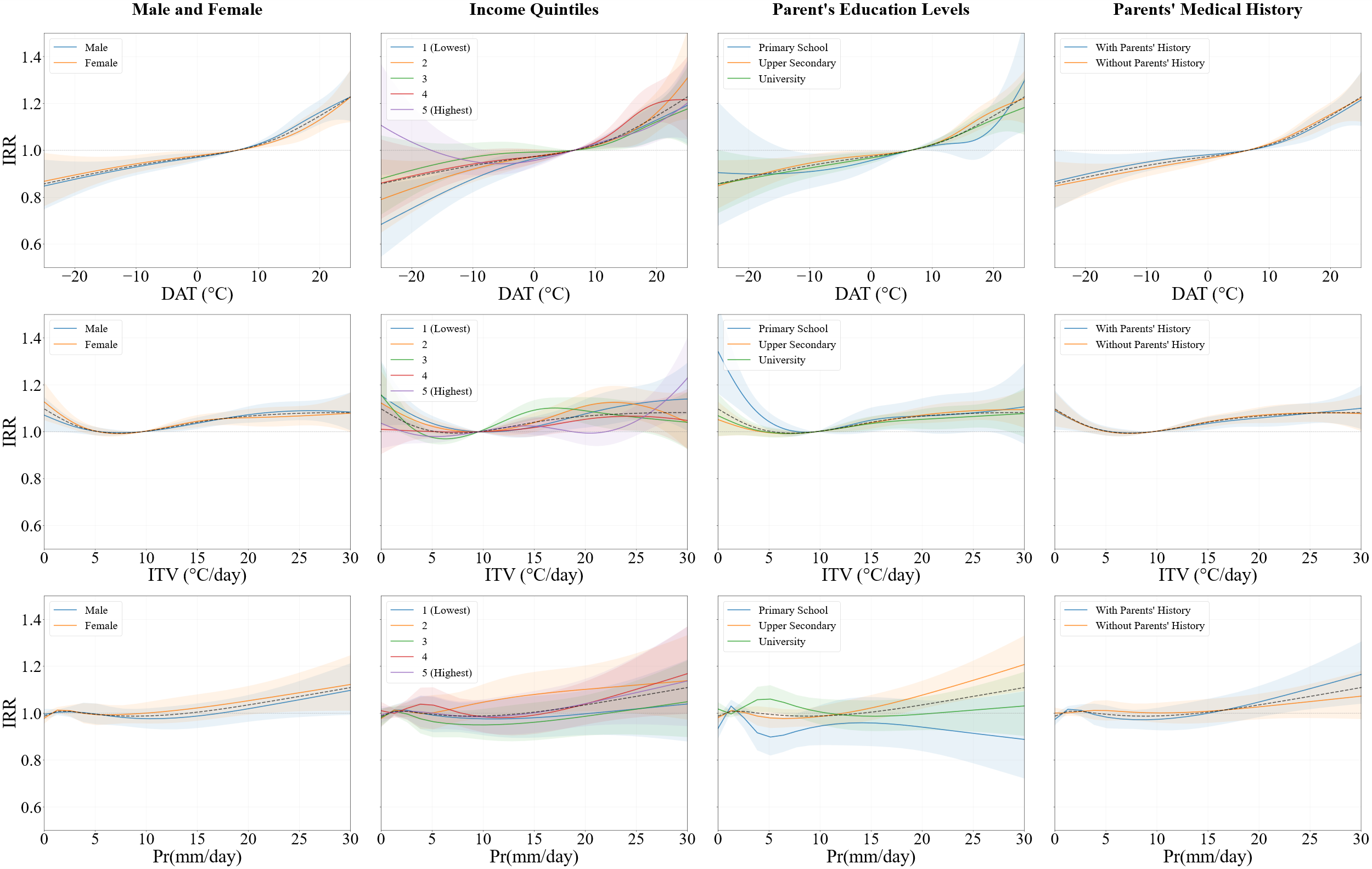

Figure S12: Stratified exposure-response associations for all-cause hospitalizations. IRRs are shown across Daily Average Temperature, Intraday Temperature Variability and Precipitation (top to bottom) by sex, income quintile, parent’s educational levels, and parent’s medical history

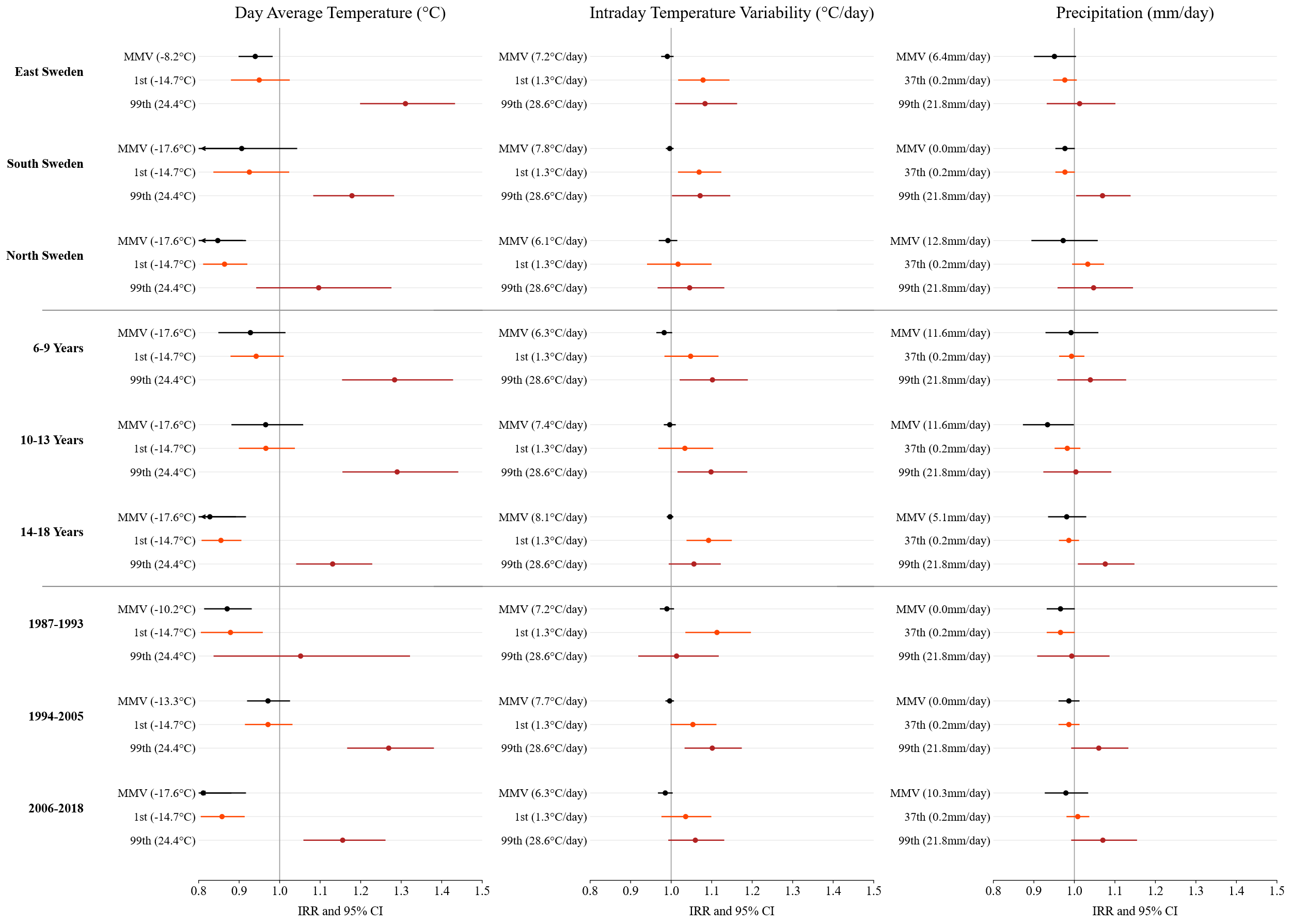

Figure S13: Stratified exposure-response associations for all-cause hospitalizations. IRRs are shown across different climate variables by region (landsdelar, age group, and calendar period, for 1st and 99th percentile values of Daily Average Temperature, Intraday Temperature Variability and Precipitation (top to bottom).

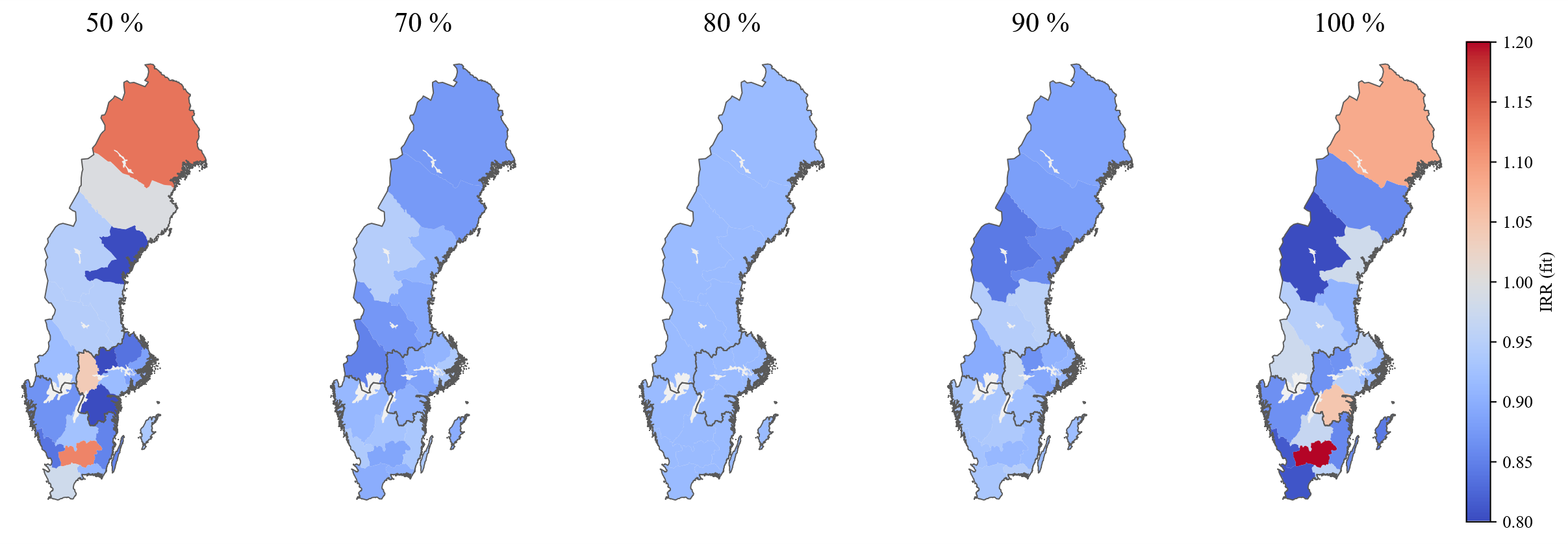

Figure S14: Swedish counties map for different intervals of Relative Humidity

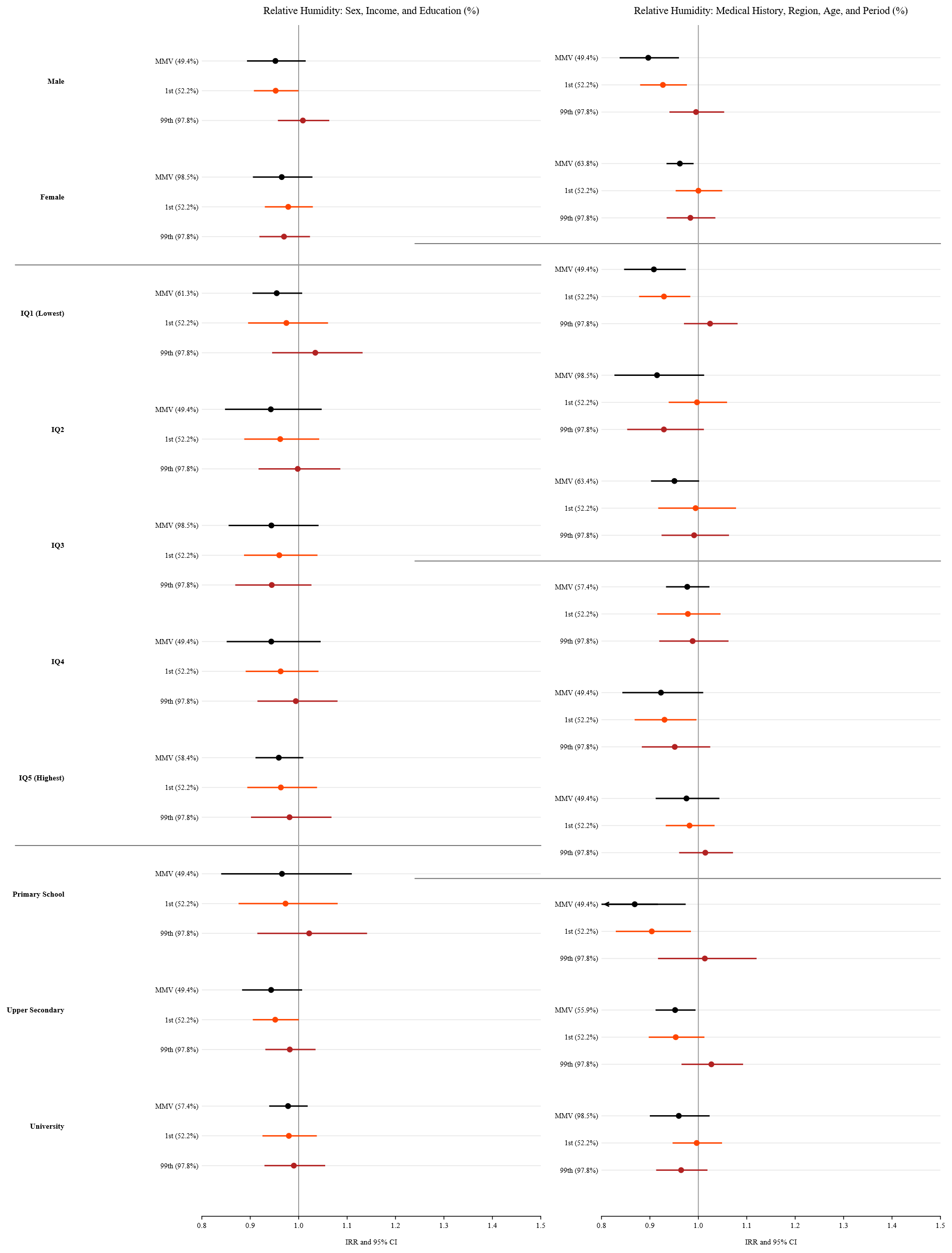

Figure S15: Stratified exposure-response associations for all-cause hospitalizations. IRRs are shown by sex, age group and parental education for 1^st^, 99^th^ percentile and most optimal values of Relative Humidity (%).

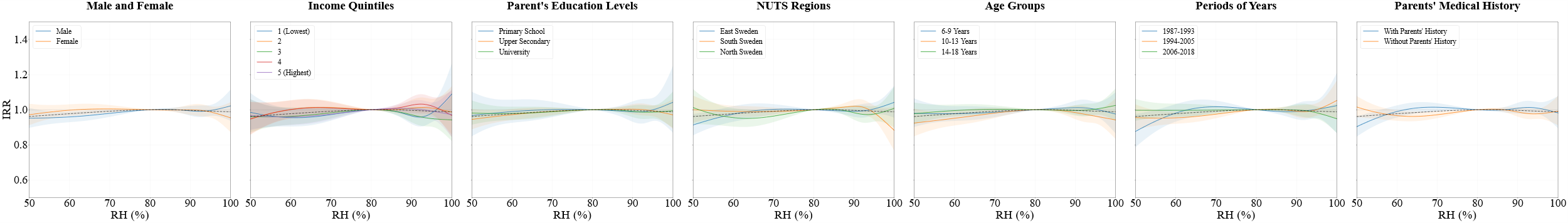

Figure S16: Stratified exposure-response associations for all-cause hospitalizations. IRRs are shown for Relative Humidity (%) by sex, income quintiles, parents’ education age group, calendar period, region and individuals with parental medical history

Table S4: Supplementary Table SX. Wald tests for effect modification of the cumulative exposure–lag–response association (lag 0–21 days). Values are χ² (df); p-value.

| **Modifier** | **DAT** | **ITV** | **RH** | **Pr** |
| --- | --- | --- | --- | --- |
| Sex | 72·5 (20); <0.001 | 82·9 (20); <0.001 | 25·8 (20); 0.174 | 26·8 (20); 0·142 |
| Age group | 132·8 (40); <0·001 | 158·8 (40); <0·001 | 45·5 (40); 0·253 | 48·8 (40); 0·161 |
| Season | 331·8 (20); <0·001 | 142·0 (20); <0·001 | 304·8 (20); <0·001 | 97·6 (20); <0·001 |
| Calendar period | 757·2 (40); <0·001 | 659·6 (40); <0·001 | 369·9 (40); <0·001 | 154·6 (40); <0·001 |
| Region (NUTS1) | 185·0 (40); <0·001 | 71·3 (40); 0·002 | 318·7 (40); <0·001 | 159·2 (40); <0·001 |
| Parental income quintile | 84·7 (80); 0·337 | 106·2 (80); 0·027 | 71·6 (80); 0·736 | 109·5 (80); 0·016 |
| Parental education | 55·4 (40); 0·053 | 56·4 (40); 0·044 | 36·1 (40); 0·644 | 52·1 (40); 0·095 |
| Parental medical history | 35·6 (20); 0·017 | 21·4 (20); 0·375 | 27·7 (20); 0·116 | 20·3 (20); 0·442 |
| Acute outcome | 142·5 (20); <0·001 | 57·7 (20); <0·001 | 19·2 (20); 0·509 | 17·9 (20); 0·593 |
| Chronic outcome | 103·1 (20); <0·001 | 43·4 (20); 0·002 | 57·0 (20); <0·001 | 36·1 (20); 0·015 |
| Cause of admission (5 categories) | 958·2 (80); <0·001 | 1236·0 (80); <0·001 | 196·4 (80); <0·001 | 154·7 (80); <0·001 |

### References

1	Utbildningsdepartementet. Skollag. Sveriges Riksdag, 2010 https://www.riksdagen.se/sv/dokument-lagar/dokument/svensk-forfattningssamling/skollag-2010800_sfs-2010-800.

2	Hallsén S. The Rise of Supplementary Education in Sweden: Arguments, Thought Styles, and Policy Enactment. *ECNU Review of Education* 2021; **4**: 476–93.

3	European Union. Regions in the European Union Nomenclature of territorial units for statistics (NUTS) 2024 edition. *Publications Office of the European Union* 2024. DOI:10.2785/714519.

4	Sidenvall E. Church and State in Sweden: A Contemporary Report. *Kirchliche Zeitgeschichte* 2012; **25**: 311–9.

5	REGINA - Uttagssystemet för regionala indelningar. 2025. https://regina.scb.se/ (accessed Sept 30, 2025).

6	Finansdepartementet S3. Förordning om distrikt. Sveriges Riksdag, 2015 https://www.riksdagen.se/sv/dokument-och-lagar/dokument/svensk-forfattningssamling/forordning-2015493-om-distrikt_sfs-2015-493/ (accessed Sept 30, 2025).

7	Tol RSJ. Population and trends in the global mean temperature. *Atmósfera* 2017; **30**: 121–35.

8	Andersson S, Bärring L, Landelius T, Samuelsson P, Schimanke S. SMHI Gridded Climatology. 2021.

9	Gasparrini A. Modelling lagged associations in environmental time series data: a simulation study. *Epidemiology* 2016; **27**: 835.

10	Gasparrini A. The Case Time Series Design. *Epidemiology* 2021; **32**: 829–37.

11	Gasparrini A, Armstrong B, Kenward MG. Distributed lag non-linear models. *Stat Med* 2010; **29**: 2224–34.

12	Armstrong BG, Gasparrini A, Tobias A. Conditional Poisson models: a flexible alternative to conditional logistic case cross-over analysis. *BMC Med Res Methodol* 2014; **14**: 122.

13	Gasparrini A. A tutorial on the case time series design for small-area analysis. *BMC Medical Research Methodology 2022 22:1* 2022; **22**: 129-.

14	Gasparrini A, Masselot P, Scortichini M, *et al.* Small-area assessment of temperature-related mortality risks in England and Wales: a case time series analysis. *Lancet Planet Health* 2022; **6**: e557–64.

### Table: The RECORD statement – checklist of items, extended from the STROBE statement

|  | Item No. | STROBE items | Location in manuscript | RECORD items | Location in manuscript |
| --- | --- | --- | --- | --- | --- |
| Title and abstract | | | | | |
|  | 1 | (a) Indicate the study’s design with a commonly used term in the title or the abstract (b) Provide in the abstract an informative and balanced summary of what was done and what was found | Page 1 | RECORD 1.1: The type of data used should be specified in the title or abstract. When possible, the name of the databases used should be included.  RECORD 1.2: If applicable, the geographic region and timeframe within which the study took place should be reported in the title or abstract.  RECORD 1.3: If linkage between databases was conducted for the study, this should be clearly stated in the title or abstract. | Page 1 |
| Background rationale | 2 | Explain the scientific background and rationale for the investigation being reported | Page 3 |  |  |
| Objectives | 3 | State specific objectives, including any prespecified hypotheses | Page 3 |  |  |
| Methods | | | | | |
| Study Design | 4 | Present key elements of study design early in the paper | Page 3-4  Supplementary Methods |  |  |
| Setting | 5 | Describe the setting, locations, and relevant dates, including periods of recruitment, exposure, follow-up, and data collection | Page 3-4  Supplementary Methods  Table S1 |  |  |
| Participants | 6 | (a) Cohort study - Give the eligibility criteria, and the sources and methods of selection of participants. Describe methods of follow-up  Case-control study - Give the eligibility criteria, and the sources and methods of case ascertainment and control selection. Give the rationale for the choice of cases and controls  Cross-sectional study - Give the eligibility criteria, and the sources and methods of selection of participants  (b) Cohort study - For matched studies, give matching criteria and number of exposed and unexposed  Case-control study - For matched studies, give matching criteria and the number of controls per case | Page 3-4  Supplementary Methods | RECORD 6.1: The methods of study population selection (such as codes or algorithms used to identify subjects) should be listed in detail. If this is not possible, an explanation should be provided.  RECORD 6.2: Any validation studies of the codes or algorithms used to select the population should be referenced. If validation was conducted for this study and not published elsewhere, detailed methods and results should be provided.  RECORD 6.3: If the study involved linkage of databases, consider use of a flow diagram or other graphical display to demonstrate the data linkage process, including the number of individuals with linked data at each stage. | Page 3-4  Supplementary Methods  Table 1 and Table S2 |
| Variables | 7 | Clearly define all outcomes, exposures, predictors, potential confounders, and effect modifiers. Give diagnostic criteria, if applicable. | Page 4  Supplementary Methods | RECORD 7.1: A complete list of codes and algorithms used to classify exposures, outcomes, confounders, and effect modifiers should be provided. If these cannot be reported, an explanation should be provided. | Page 4  Supplementary Methods  Table 1  Table S2 |
| Data sources/ measurement | 8 | For each variable of interest, give sources of data and details of methods of assessment (measurement).  Describe comparability of assessment methods if there is more than one group | Page 4  Supplementary Methods |  |  |
| Bias | 9 | Describe any efforts to address potential sources of bias | Page 4-5  Table S1 |  |  |
| Study size | 10 | Explain how the study size was arrived at | Page 6  Table 1  Table S1 |  |  |
| Quantitative variables | 11 | Explain how quantitative variables were handled in the analyses. If applicable, describe which groupings were chosen, and why | Page 4-5  Supplementary Methods |  |  |
| Statistical methods | 12 | (a) Describe all statistical methods, including those used to control for confounding  (b) Describe any methods used to examine subgroups and interactions  (c) Explain how missing data were addressed  (d) Cohort study - If applicable, explain how loss to follow-up was addressed  Case-control study - If applicable, explain how matching of cases and controls was addressed  Cross-sectional study - If applicable, describe analytical methods taking account of sampling strategy  (e) Describe any sensitivity analyses | Page 4-5  Supplementary Methods |  |  |
| Data access and cleaning methods |  |  |  | RECORD 12.1: Authors should describe the extent to which the investigators had access to the database population used to create the study population.  RECORD 12.2: Authors should provide information on the data cleaning methods used in the study. | Page 3-6  Supplementary Methods |
| Linkage |  | .. |  | RECORD 12.3: State whether the study included person-level, institutional-level, or other data linkage across two or more databases. The methods of linkage and methods of linkage quality evaluation should be provided. | Page 3  Supplementary Methods |
| Results | | | | | |
| Participants | 13 | (a) Report the numbers of individuals at each stage of the study (e.g., numbers potentially eligible, examined for eligibility, confirmed eligible, included in the study, completing follow-up, and analysed)  (b) Give reasons for non-participation at each stage.  (c) Consider use of a flow diagram | Table 1  Table S2  Pages 6-7  Supplementary Methods | RECORD 13.1: Describe in detail the selection of the persons included in the study (i.e., study population selection) including filtering based on data quality, data availability and linkage. The selection of included persons can be described in the text and/or by means of the study flow diagram. | Table 1  Pages 6-7 |
| Descriptive data | 14 | (a) Give characteristics of study participants (e.g., demographic, clinical, social) and information on exposures and potential confounders  (b) Indicate the number of participants with missing data for each variable of interest  (c) Cohort study - summarise follow-up time (e.g., average and total amount) | Table 1  Pages 6 |  |  |
| Outcome data | 15 | Cohort study - Report numbers of outcome events or summary measures over time  Case-control study - Report numbers in each exposure category, or summary measures of exposure  Cross-sectional study - Report numbers of outcome events or summary measures | Table S1  Supplementary Methods  Page 6 |  |  |
| Main results | 16 | (a) Give unadjusted estimates and, if applicable, confounder-adjusted estimates and their precision (e.g., 95% confidence interval). Make clear which confounders were adjusted for and why they were included  (b) Report category boundaries when continuous variables were categorized  (c) If relevant, consider translating estimates of relative risk into absolute risk for a meaningful time period | Page 6-10  Figure 1-5 |  |  |
| Other analyses | 17 | Report other analyses done—e.g., analyses of subgroups and interactions, and sensitivity analyses | Supplementary Methods  Figure S1-S12 |  |  |
| Discussion | | | | | |
| Key results | 18 | Summarise key results with reference to study objectives | Page 10-11 |  |  |
| Limitations | 19 | Discuss limitations of the study, taking into account sources of potential bias or imprecision. Discuss both direction and magnitude of any potential bias | Page 11 | RECORD 19.1: Discuss the implications of using data that were not created or collected to answer the specific research question(s). Include discussion of misclassification bias, unmeasured confounding, missing data, and changing eligibility over time, as they pertain to the study being reported. | Page 12 |
| Interpretation | 20 | Give a cautious overall interpretation of results considering objectives, limitations, multiplicity of analyses, results from similar studies, and other relevant evidence | Page 11 |  |  |
| Generalisability | 21 | Discuss the generalisability (external validity) of the study results | Page 12 |  |  |
| Other Information | | | | | |
| Funding | 22 | Give the source of funding and the role of the funders for the present study and, if applicable, for the original study on which the present article is based |  |  | Page 6 |
| Accessibility of protocol, raw data, and programming code |  |  |  | RECORD 22.1: Authors should provide information on how to access any supplemental information such as the study protocol, raw data, or programming code. | Pages 6 and 12  Github repository |
